# The ITE-Transformer: A pre-trained foundation model for predicting individualized treatment effects from electronic health records

**DOI:** 10.64898/2026.11.19.26368479

**Authors:** Clemente Arceri, Emmanouil Angelis, Philip Heesen, Valentyn Melnychuk, Martin Bendszus, Stefan Bauer, Stefan Feuerriegel

## Abstract

Transformer-based foundation models have shown strong potential for modeling longitudinal electronic health records (EHRs), yet existing models focus primarily on risk prediction rather than treatment effect estimation, which limits evidence generation. Here, we introduce the ITE-Transformer, a transformer-based framework that combines self-supervised representation learning from longitudinal patient trajectories with doubly robust causal machine learning for predicting individualized treatment effects (ITEs). As a result, our ITE-Transformer allows to study counterfactual questions of how an individual patient’s outcome may differ under alternative treatment strategies. The ITE-Transformer learns temporally contextualized representations of patients’ diagnosis and medication histories and subsequently uses these representations to predict treatment assignment, treatment-specific outcome risks, and patient-specific ITEs. The ITE-Transformer is trained using structured EHR data from *∼*350,000 participants in the *All of Us Research Program* and evaluated in two clinically different comparative-effectiveness tasks: (i) GLP-1 receptor agonists versus DPP-4 inhibitors among patients with type 2 diabetes, with major adverse cardiovascular events as the outcome, and (ii) immunotherapy versus chemotherapy in a pan-cancer cohort, with all-cause mortality as the outcome. Across both applications, the ITE-Transformer improves the discrimination and calibration of treatment assignment and outcome models compared with existing tabular machine learning approaches and an otherwise identical transformer trained without self-supervised pre-training. The ITE-Transformer further identifies substantial heterogeneity in predicted ITEs and yields interpretable patterns of treatment response associated with patients’ diagnoses, medication histories, and comorbidities. Together, our work demonstrates how clinical foundation models can be developed for analyzing heterogeneity in treatment effects and thus support real-world evidence generation.

## Introduction

Estimating individualized treatment effects (ITEs) provides the evidence base for precision medicine by helping clinicians identify patients most likely to benefit from a given treatment [1–3]. Longitudinal electronic health records (EHRs) offer a rich source of real-world data for this purpose, capturing patients’ diagnoses, treatments, and disease progression over time [4–6]. Here, we present ITE-Transformer, a pre-trained clinical foundation model that combines longitudinal patient representations and causal machine learning [2] to predict ITEs from EHR data.

Recent advances in transformer-based foundation models have demonstrated strong performance in modeling longitudinal patient trajectories from EHR data by capturing complex dependencies across diagnoses, treatments, and disease progression [7–14]. However, existing clinical foundation models have largely focused on disease risk prediction (e.g., Delphi-2M predicts the risk of over 1,000 diseases [10]). While extremely valuable for risk stratification and preventive care, such foundation models do not directly provide comparative clinical evidence on how patients would respond to different treatments. Estimating such treatment effects involves counterfactual questions and, when using real-world data, requires accounting for non-random treatment assignment and confounding [15–17]. Importantly, failing to account for such confounders can lead to biased estimates, as patients receiving more intensive treatments may also have more severe disease and worse underlying prognoses. In principle, causal machine learning [2, 18] offers methods for estimating treatment effects from high-dimensional data [2], yet approaches specifically tailored to clinical foundation models are missing.

We introduce ITE-Transformer, a pre-trained transformer-based foundation model that combines longitudinal EHR representations with doubly robust causal machine learning to predict ITEs. The ITE-Transformer is pre-trained on temporally ordered sequences of clinical events to learn contextualized representations of patient histories; these representations are subsequently used to predict treatment assignment and treatment-specific outcome risks, which are combined within a doubly robust framework to compute ITEs. The ITE-Transformer is pre-trained on structured EHR data from *∼*350,000 patients in the *All of Us* Research Program [19]. We evaluate the ITE-Transformer using two clinically different treatment comparisons: (i) GLP-1 receptor agonists versus DPP-4 inhibitors in patients with type 2 diabetes, and (ii) immunotherapy versus chemotherapy in a pan-cancer cohort spanning multiple malignant tumor types. More broadly, this work extends clinical foundation models beyond risk prediction toward analyzing ITEs for evidence generation.

## Results

### A foundation model for ITE prediction

We developed the ITE-Transformer, a transformer-based foundation model for predicting ITEs from longitudinal electronic health records (EHRs). We pre-trained the ITE-Transformer on structured EHRs from approximately 350,000 participants enrolled in the *All of Us Research Program* [19], a large, demographically diverse longitudinal cohort in the United States. The ITE-Transformer used patient trajectories as input, including demographic characteristics, diagnosis codes, and medication exposures. Diagnoses were encoded using level-3 ICD codes from chapters I–XVII; medications were encoded using level 4 Anatomical Therapeutic Chemical (ATC) codes. Detailed cohort characteristics are provided in Supplementary Table S1 and Supplementary Figure S1.

Patient histories were represented as temporally ordered sequences of clinical events (Figure 1a), analogous to token sequences in language modeling. Tokens were ordered by event time and paired with the patient’s age at the corresponding event. A token indicating biological sex was added at the beginning of each sequence. We used a standard decoder-only transformer architecture with causal self-attention to ensure that each token attended only to preceding clinical events and thus to prevent tokens from attending to subsequent events in the patient’s trajectory. The architecture was customized for the clinical setting by replacing conventional positional encoding with a continuous age-based encoding [10] to capture the irregular timing of patient trajectories. The ITE-Transformer was then pre-trained using a self-supervised next-token prediction objective (Figure 1b).

**Figure 1:**
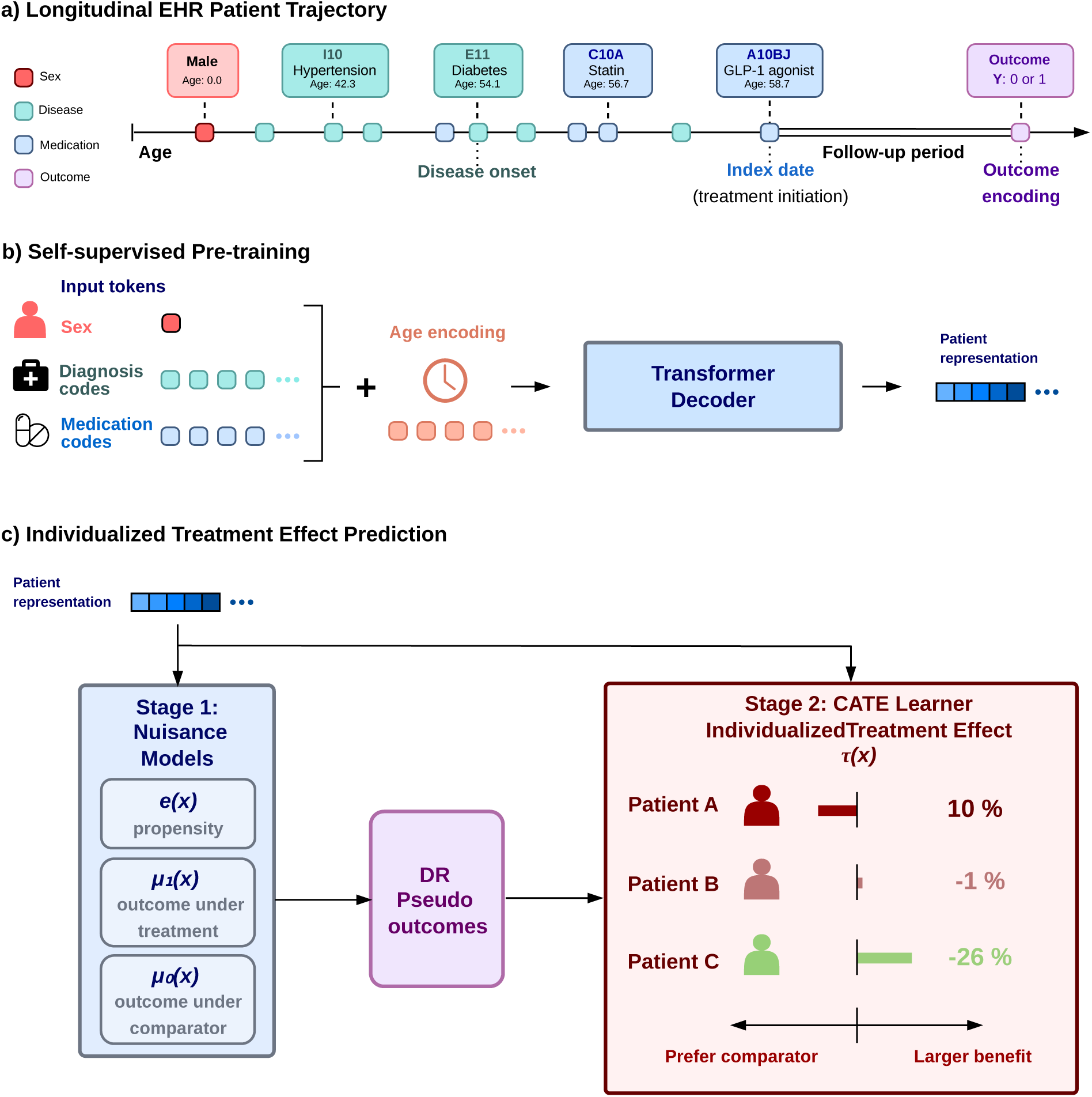
Overview of the ITE-Transformer. **a**, Example patient trajectory represented as temporally ordered sequence of clinical events, including diagnoses and medication exposures. Pre-treatment clinical histories serve as model inputs, while outcomes are assessed during post-treatment follow-up (i.e., after the index date). **b**, Self-supervised pre-training of the decoder-only transformer on longitudinal EHR trajectories using next-token prediction. Clinical tokens are combined with age-based temporal encodings. Details about the architecture are in Supplementary Figure S2. **c**, ITE-Transformer predicts the individualized treatment effects (ITEs), defined by the conditional average treatment effect. ITE-Transformer uses a doubly robust (DR) approach from causal machine learning [18,20,21] to predict the ITEs. In the first stage, transformer-based prediction heads estimate treatment assignment probabilities and treatment-specific outcome risks, which are combined with observed treatments and outcomes to construct doubly robust pseudo-outcomes using augmented inverse probability weighting (AIPW). In the second stage, a conditional average treatment effect (CATE) learner uses these pseudo-outcomes to compute the ITEs.

For downstream ITE prediction (Figure 1c), we integrated the pre-trained transformer into a causal machine learning framework that builds on principles of doubly robust estimation [18, 20, 21]. The idea of doubly robust learners is to, instead of estimating the treatment effects directly, to first model both the treatment assignment and the treatment-specific outcomes separately, before combining these predictions to estimate treatment effects in a way that reduces sensitivity to misspecification of either prediction component. Importantly, both stages leverage the longitudinal patient representations learned by the pre-trained transformer. For this, we use the hidden representation of the last non-padded token before the treatment index date to summarize each patient’s pre-treatment clinical history. Formally, the ITE-Transformer proceeds in two stages (Figure 1c). In the first stage, dedicated prediction heads are attached to the transformer to jointly estimate two nuisance components (i.e., auxiliary predictions that are used in the second stage), namely, the propensity for treatment assignment and the expected outcomes under each treatment alternative. The estimates are then used to construct doubly robust pseudo-outcomes based on augmented inverse probability weighting (AIPW) [18]. AIPW combines outcome predictions with inverse-propensity-weighted corrections based on observed treatments and outcomes, thus adjusting for measured differences in treatment assignment, while reducing sensitivity to errors in the nuisance components. In the second stage, a dedicated prediction head is used to learn the conditional average treatment effect (CATE) from the pseudo-outcomes using the transformer-derived patient representations, while keeping the transformer backbone frozen. As a result, the output are patient-specific CATE predictions, which we refer to as ITEs and which predict the expected difference in outcome risk between the study treatment and the comparator varies across patients with different pre-treatment histories.

We evaluated the ITE-Transformer across two clinically different comparative-effectiveness tasks: GLP-1 receptor agonists versus DPP-4 inhibitors in patients with type 2 diabetes, and immunotherapy versus chemotherapy in a pan-cancer cohort. For both tasks, we used patients’ pre-treatment clinical histories before treatment initiation as model inputs and assessed outcomes during post-treatment follow-up.

### Learned representations of medical concepts

The ITE-Transformer learned embeddings that reflect clinically meaningful relationships among medical concepts. To demonstrate this, we projected the high-dimensional learned embeddings onto a two-dimensional visualization using uniform manifold approximation and projection (UMAP) [22]. In the UMAP projection (Figure 2), diagnosis and medication concepts formed largely separated regions, despite being trained jointly without imposing modality-specific constraints. Within these regions, concepts belonging to related ICD and ATC chapters tended to cluster together, suggesting that the learned embeddings align with the high-level structure of the clinical coding systems.

**Figure 2:**
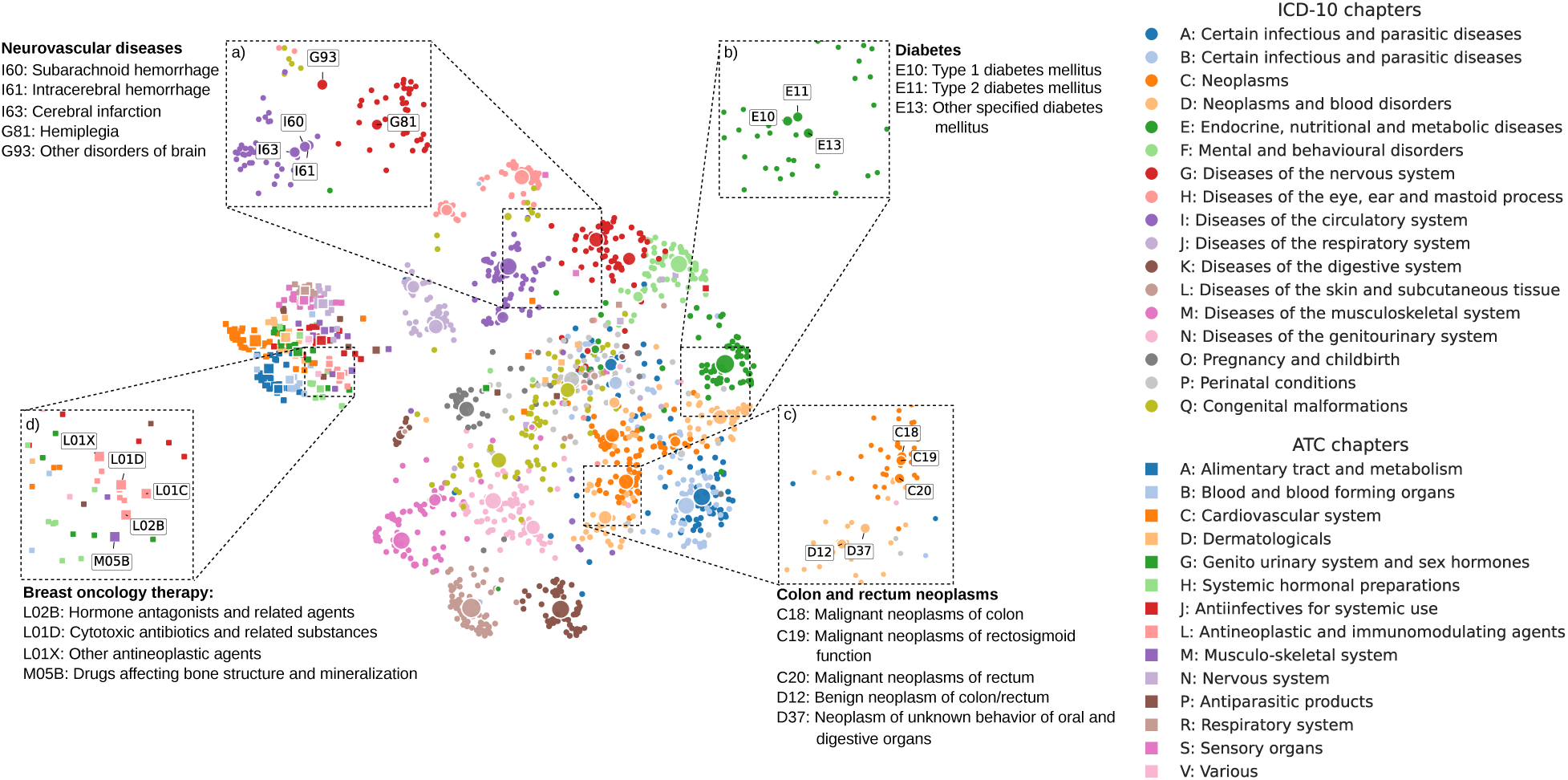
Learned embeddings of the ITE-Transformer. UMAP projection of the learned clinical token embeddings after pre-training. Each point represents a diagnosis, while squares represent medication codes, with colors indicating the respective ICD-10 and ATC chapters. Related clinical concepts cluster together, while diagnoses and medications form largely distinct regions. Enlarged views highlight several examples for: **a**, neurovascular diseases; **b**, diabetes; **c**, colon and rectum neoplasms; and **d**, breast oncology therapy. These patterns demonstrate how the learned embeddings capture clinical relationships both within and across established coding hierarchies.

The embeddings also capture more fine-grained clinical structure. For example, related diagnoses, such as diabetes codes (Figure 2b) and anatomically-related neoplasm codes (Figure 2c), clustered closely together. Codes representing clinically-related conditions but from different ICD chapters appeared also in close proximity, such as illustrated by conditions affecting the blood vessels supplying the brain (Figure 2a). This suggests that the learned embeddings capture clinical relationships beyond predefined coding hierarchies. Overall, these findings show that the ITE-Transformer learns clinically coherent representations of medical concepts, thereby providing a foundation for contextualized representations of patient trajectories.

### The ITE-Transformer learns clinical patterns across treatments and outcomes for ITE prediction

To assess whether ITE-Transformer captures clinically relevant information for predicting ITEs, we evaluated how well the ITE-Transformer-based nuisance components predict treatment assignment and clinical outcomes. For this, we use two cohorts from the *All of Us Research Program*. The type 2 diabetes cohort included 8,540 patients receiving GLP-1 receptor agonists or DPP-4 inhibitors, with major adverse cardiovascular events (MACE) assessed within three years after treatment initiation. The pan-cancer cohort included 4,998 patients receiving immunotherapy or chemotherapy, with all-cause mortality assessed again over a three-year period. Cohort characteristics are summarized in Table 1; detailed eligibility criteria are provided in the Methods section.

**Table 1:** Cohort characteristics for comparative-effectiveness analyses. Outcome rates represent the proportion of patients experiencing major adverse cardiovascular events (MACE) in the diabetes cohort or all-cause mortality in the pan-cancer cohort within three years after treatment initiation, pooled across treatment groups.

|  | **GLP-1 vs DPP-4** | **Immuno- vs chemotherapy** |
|---|---|---|
| Patients | 8,540 | 4,998 |
| Treated / comparator | 3,600 / 4,940 | 1,008 / 3,990 |
| Outcome | MACE | Death |
| Outcome rate | 14.2% | 6.9% |

Because counterfactual outcomes are not observed in real-world data, ITE predictions cannot be evaluated directly against ground-truth observations [3]. Following best practice in causal machine learning [2, 23], we therefore evaluated the first-stage nuisance components underlying treatment effect estimation; i.e., the component predicting the propensity of treatment assignment and the outcome component predicting factual outcome risk under observed treatment choice. We compared the ITE-Transformer against several baseline models from prior literature [10], including an age-and-sex classifier, a random forest, XGBoost, and an otherwise identical transformer trained on the downstream task without self-supervised pre-training (which we refer to as NP-Transformer). We then assessed model discrimination using the area under the receiver operating characteristic curve (AUROC) and the area under the precision–recall curve (AUPRC). We also assessed model calibration comparing predicted treatment assignment probabilities and outcome risks with their corresponding observed frequencies across probability bins. Together, these analyses assess the reliability of the nuisance components used for downstream ITE prediction.

Across both comparisons, the ITE-Transformer consistently outperformed the baselines in predicting both treatment propensity and factual outcomes. In the GLP-1 versus DPP-4 comparison (Figure 3a,c), the ITE-Transformer improved over the strongest baseline by 1.6% in AUROC and 1.5% in AUPRC in treatment propensity prediction, and by 1.2% in AUROC and 2.1% in AUPRC in outcome risk prediction. In the pan-cancer immunotherapy versus chemotherapy comparison (Figure 4a,c), the corresponding improvements were 2.1% in AUROC and 2.7% in AUPRC in treatment propensity prediction, and by 0.8% in AUROC and 0.5% in AUPRC in outcome risk prediction. These improvements suggest that self-supervised pre-training on longitudinal EHR trajectories yields more informative patient representations for the predictive models underlying ITE prediction.

**Figure 3:**
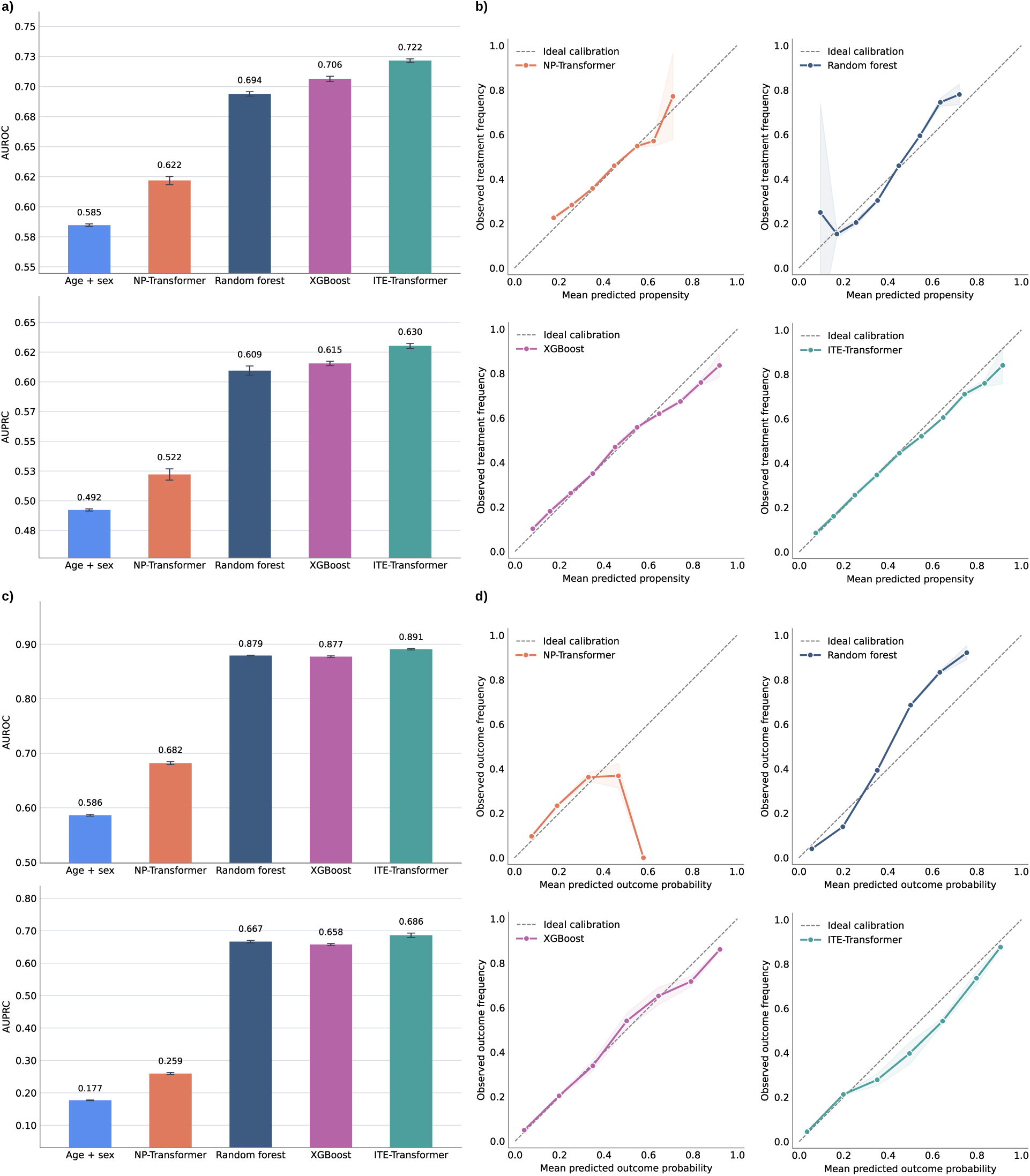
Performance of the nuisance components for the GLP-1 receptor agonist versus DPP-4 inhibitor comparison. **a**, Discriminative performance of the propensity model predicting treatment assignment from pre-treatment patient histories. **b**, Calibration of the propensity model. **c**, Discriminative performance of the outcome models predicting major adverse cardiovascular events (MACE) conditional on patient history and treatment assignment. **d**, Calibration of the outcome models. Discrimination is assessed using the area under the receiver operating characteristic curve (AUROC) and the area under the precision–recall curve (AUPRC). Calibration compares predicted probabilities with observed frequencies across probability bins, with the diagonal indicating perfect calibration. Whiskers and shaded areas show 95% confidence intervals across five random seeds.

**Figure 4:**
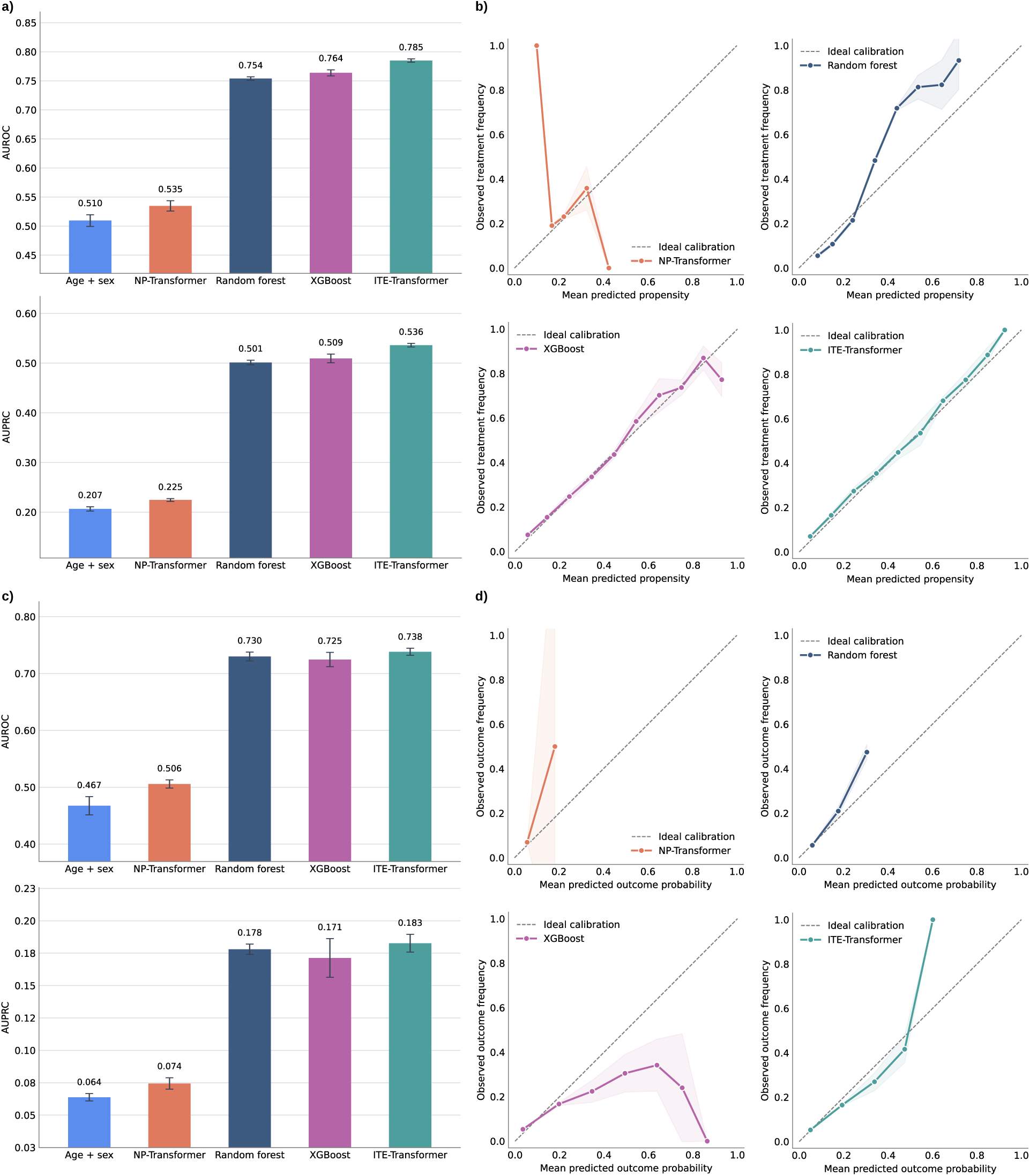
Performance of the nuisance components for the immunotherapy versus chemotherapy comparison. **a**, Discriminative performance of the propensity model predicting treatment assignment from pre-treatment patient histories. **b**, Calibration of the propensity model. **c**, Discriminative performance of the outcome models predicting three-year mortality conditional on patient history and treatment assignment. **d**, Calibration of the outcome models. Discrimination is assessed using the area under the receiver operating characteristic curve (AUROC) and the area under the precision–recall curve (AUPRC). Calibration compares predicted probabilities with observed frequencies across probability bins, with the diagonal indicating perfect calibration. Whiskers and shaded areas show 95% confidence intervals across five random seeds.

We next assessed the calibration of the nuisance components for predicting both treatment assignment propensity and factual outcome risk (see the calibration curves in Figure 3b,d for the GLP-1 versus DPP-4 comparison and Figure 4b,d for the pan-cancer comparison). Across both comparisons, the ITE-Transformer showed better calibration than the baselines, with predicted treatment assignment probabilities and outcome risks more closely matching the observed proportions of patients receiving the study treatment and experiencing the clinical outcome, respectively. These findings indicate that the pretrained longitudinal representation not only improved discrimination but also the calibration of the predictive components for ITEs.

To assess the contribution of self-supervised pre-training, we compared the ITE-Transformer with an otherwise identical transformer trained from scratch without pre-training (i.e., the NP-Transformer). As shown by representative learning curves (Supplementary Figure S3), the pretrained ITE-Transformer converged substantially faster and reached lower validation losses for both treatment propensity and outcome prediction. In contrast, the NP-Transformer showed only gradual improvements during training. These differences are also reflected by the large improvements in predictive performance and calibration, thereby confirming that self-supervised pretraining improved the longitudinal patient representations for learning informative patient representations that support ITE prediction.

We next assessed propensity score overlap to empirically examine the positivity assumption. Predicted propensity scores showed substantial overlap between treatment groups in both cohorts, with no indication of major positivity violations (Supplementary Figure S4). Finally, we combined the cross-fitted nuisance components for treatment assignment and outcome predictions using AIPW to construct doubly robust pseudo-outcomes for training the second-stage CATE learner. The distributions of these pseudo-outcomes for both treatment comparison tasks are shown in Supplementary Figure S5.

### The ITE-Transformer identifies heterogeneity in predicted treatment response

Next, we examined how the ITEs predicted by the ITE-Transformer varied across patients. ITEs were defined as the predicted difference in adverse outcome risk under the study treatment versus the comparator, with negative values indicating a lower predicted risk under the study treatment (e.g., an ITE of *−*0.15 corresponds to a 15-percentage-point reduction in predicted risk).

Figure 5a,c shows the distributions of predicted ITEs across both comparisons. In the GLP-1 versus DPP-4 comparison, 66.0% of patients had negative predicted ITEs, indicating a lower predicted risk of MACE under GLP-1 receptor agonists than under DPP-4 inhibitors. In the pancancer comparison, 68.9% of patients had negative predicted ITEs, indicating a lower predicted risk of three-year mortality under immunotherapy than under chemotherapy. The distributions showed substantial variation in both the direction and magnitude of predicted ITEs, suggesting heterogeneity in treatment benefit across patients.

**Figure 5:**
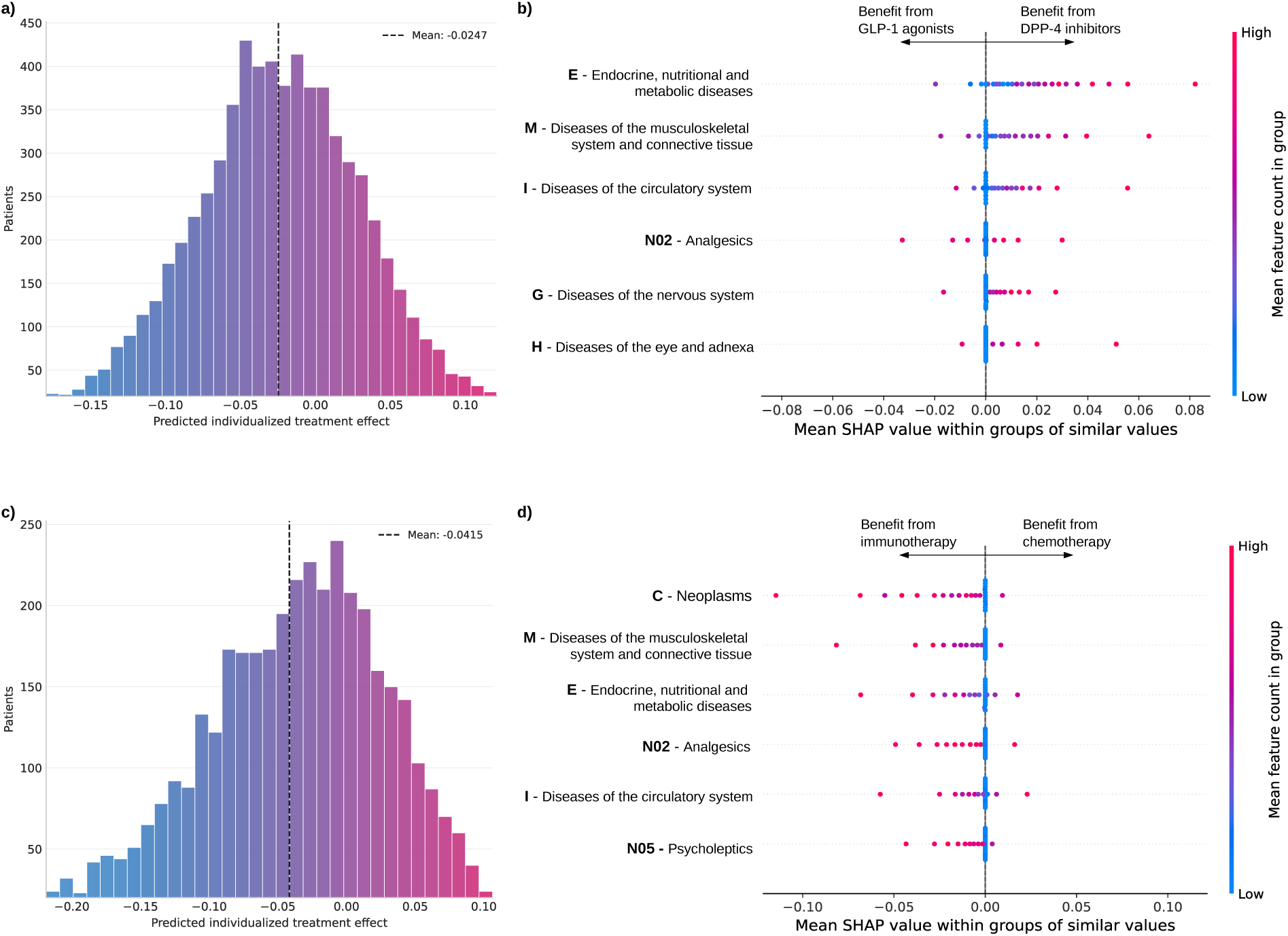
Heterogeneity in predicted ITEs and associated clinical predictors. Predicted ITEs for the two comparative-effectiveness task, namely, GLP-1 receptor agonists versus DPP-4 inhibitors and immunotherapy versus chemotherapy. Negative values indicate lower predicted risk under the study treatment (GLP-1 receptor agonists or immunotherapy), whereas positive values favor the comparator (DPP-4 inhibitors or chemotherapy). **a**, Distribution of predicted ITEs, defined as the difference in the predicted probability of MACE under GLP-1 receptor agonists versus DPP-4 inhibitors. **b**, SHAP analysis showing the diagnosis and medication groups that contribute most strongly to the predicted treatment effects. **c**, Distribution of predicted ITEs, defined as the difference in the predicted probability of death under immunotherapy versus chemotherapy. **d**, SHAP analysis showing the diagnosis and medication groups that contribute most strongly to the predicted ITEs. The magnitude of SHAP values indicates the strength of each feature’s contribution, while negative and positive values indicate shifts in predicted ITEs toward larger relative benefit from the study treatment and comparator, respectively. SHAP values are aggregated at the disease-chapter level for diagnoses and at the medication-group level for medications. To comply with dissemination regulations of the *All of Us Research Program*, the y-axis in histograms are capped at 20 patients, and the SHAP beeswarm plot shows mean SHAP values across groups of 20 patients.

We next assessed the effect score calibration [3, 24] to examine whether predicted ITEs aligned with the average treatment effects estimates across patient strata with varying treatment benefit. To this end, patients were grouped into strata according to their predicted ITEs, and the mean over the ITEs within each strata was compared with the corresponding average treatment effect estimate (Figure 6). Across both comparisons, the predicted effect scores were well calibrated and preserved the ordering across strata, suggesting that the CATE learner captured systematic variation in treatment effects.

**Figure 6:**
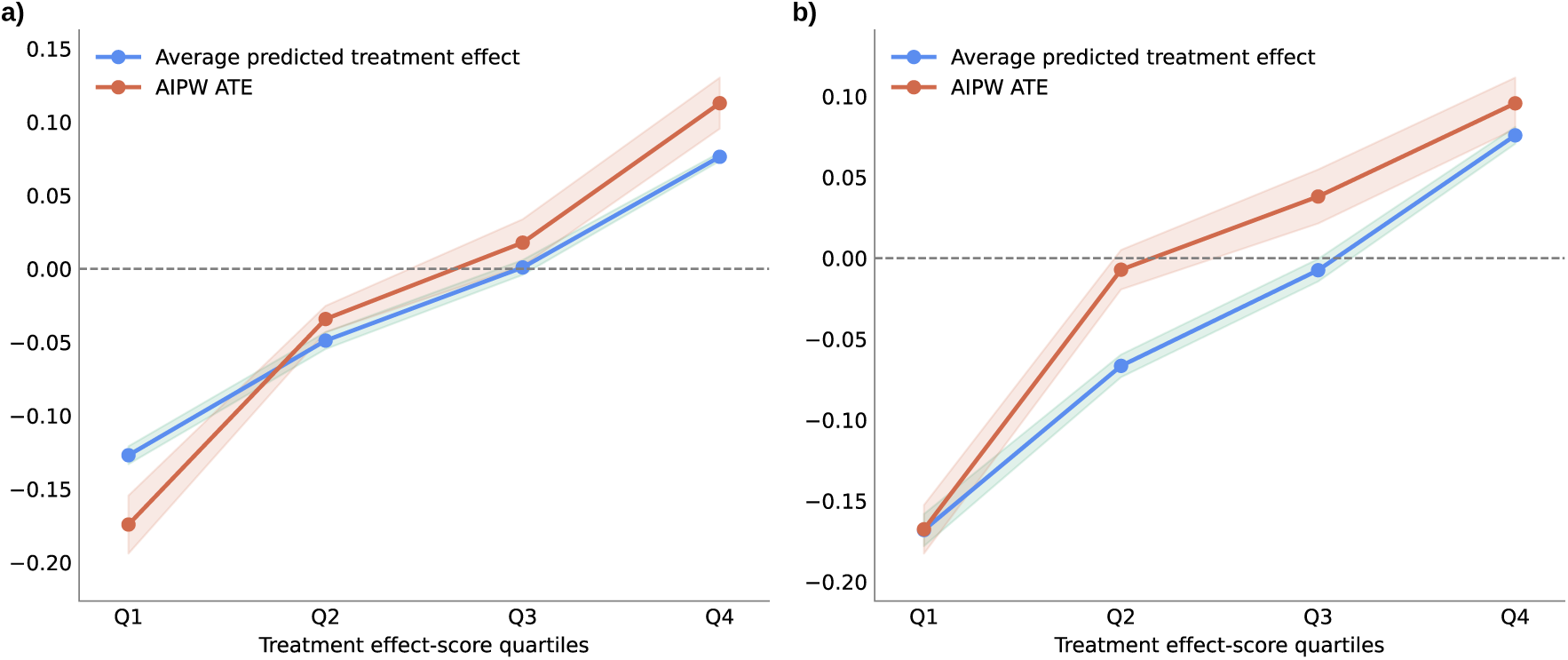
Effect score calibration. Effect score calibration [3, 24] of predicted individualized treatment effects (ITEs) for **a**, GLP-1 receptor agonist versus DPP-4 inhibitor comparison and **b**, immunotherapy versus chemotherapy comparison. For this, patients were ranked according to the ITEs predicted by the fitted CATE learner and divided into different strata (i.e., quartiles). Within each strata, the average over the predicted ITEs (in blue) was compared with the corresponding strata-specific average treatment effect (computed via cross-fitted AIPW; in red). Ideally, both effect scores should show a similar ordering across patient strata, as is the case here.

To further assess the robustness of the predicted ITEs, we compared the ITE-Transformer with a causal forest [25] trained on the same pre-trained patient representations (Supplementary Figure S6). Across both treatment comparisons, the methods showed broad agreement in group-level treatment effect estimates and their ranking, which supports the reliability of the patterns identified by the ITE-Transformer.

### The ITE-Transformer captures clinically relevant predictors of treatment effect heterogeneity

To demonstrate the interpretability of ITE-Transformer, we performed a SHAP analysis [26] to identify clinical features that contribute to the heterogeneity of the ITEs predicted by ITE-Transformer and evaluate their clinical plausibility. SHAP values quantify how strongly each feature shifts an individual prediction relative to the average predicted ITE; i.e., positive SHAP values move the prediction towards a relative benefit of the comparator, whereas negative SHAP values move it towards a relative benefit of the study treatment. To improve interpretability, individual diagnosis and medication concepts were aggregated at the ICD chapter level for diagnosis and at the ATC group level for medications.

The results are shown in Figure 5b,d. For the GLP-1 versus DPP-4 comparison, the most influential features included endocrine, nutritional, and metabolic diseases (ICD chapter E), consistent with the central role of diabetes severity, obesity, dyslipidaemia, and related metabolic disorders in determining cardiovascular risk and treatment response [27–29]. Diseases of the musculoskeletal system and connective tissue (ICD M) and the use of analgesics (ATC N02) may capture obesity-related complications, chronic pain, or reduced mobility, which are associated with cardiovascular health [30, 31]. Other influential groups included diseases of the circulatory system (ICD I), diseases of the nervous system (ICD G), and diseases of the eye and adnexa (ICD H). These features may reflect underlying cardiovascular risk, neurological comorbidities, and diabetes-related complications, thus providing additional information about patients’ overall health status.

For the immunotherapy versus chemotherapy comparison, the SHAP analysis identified clinically meaningful groups that are consistent with established determinants of cancer prognosis and treatment selection. The most influential feature group was malignant neoplasms (ICD chapter C), which captures differences in cancer type, disease burden, and disease progression [32] and are known to influence the relative effectiveness of immunotherapy and chemotherapy across different solid tumors [33, 34]. Other influential groups included musculoskeletal diseases (ICDM) and endocrine, nutritional, and metabolic diseases (ICD E). These conditions may reflect differences in comorbidity burden, body composition, functional status, and systemic inflammation, which have been linked to differences in both cancer outcomes and immune checkpoint inhibitor response [35–37].

Overall, the SHAP analysis suggests that the ITE-Transformer relies on clinically meaningful features when predicting ITEs across both clinical settings. Beyond improving model interpretability, ITE-Transformer may thus help identify patient subgroups with differential treatment benefit and help discover potential predictive biomarkers. Examining feature contributions at both the broader population level and the individual patient level may further support hypothesis generation and guide future confirmatory studies of treatment response to inform personalized treatment recommendations.

### The ITE-Transformer enables patient-level predictions of ITEs

To further illustrate how the ITE-Transformer uses longitudinal clinical histories for ITE prediction, we constructed synthetic patient trajectories representing different clinical profiles across both treatment comparisons (Figure 7). Each trajectory consisted of temporally ordered diagnosis and medication concepts, encoded using the same clinical vocabulary and age-based representation as the model inputs. We focused particularly on pairs of patients with nearly identical pre-treatment histories, which differ only in a small number of additional clinical events. This allowed us to examine how changes in specific clinical concepts may be reflected in the predicted ITEs, while keeping the remaining recorded clinical history unchanged.

**Figure 7:**
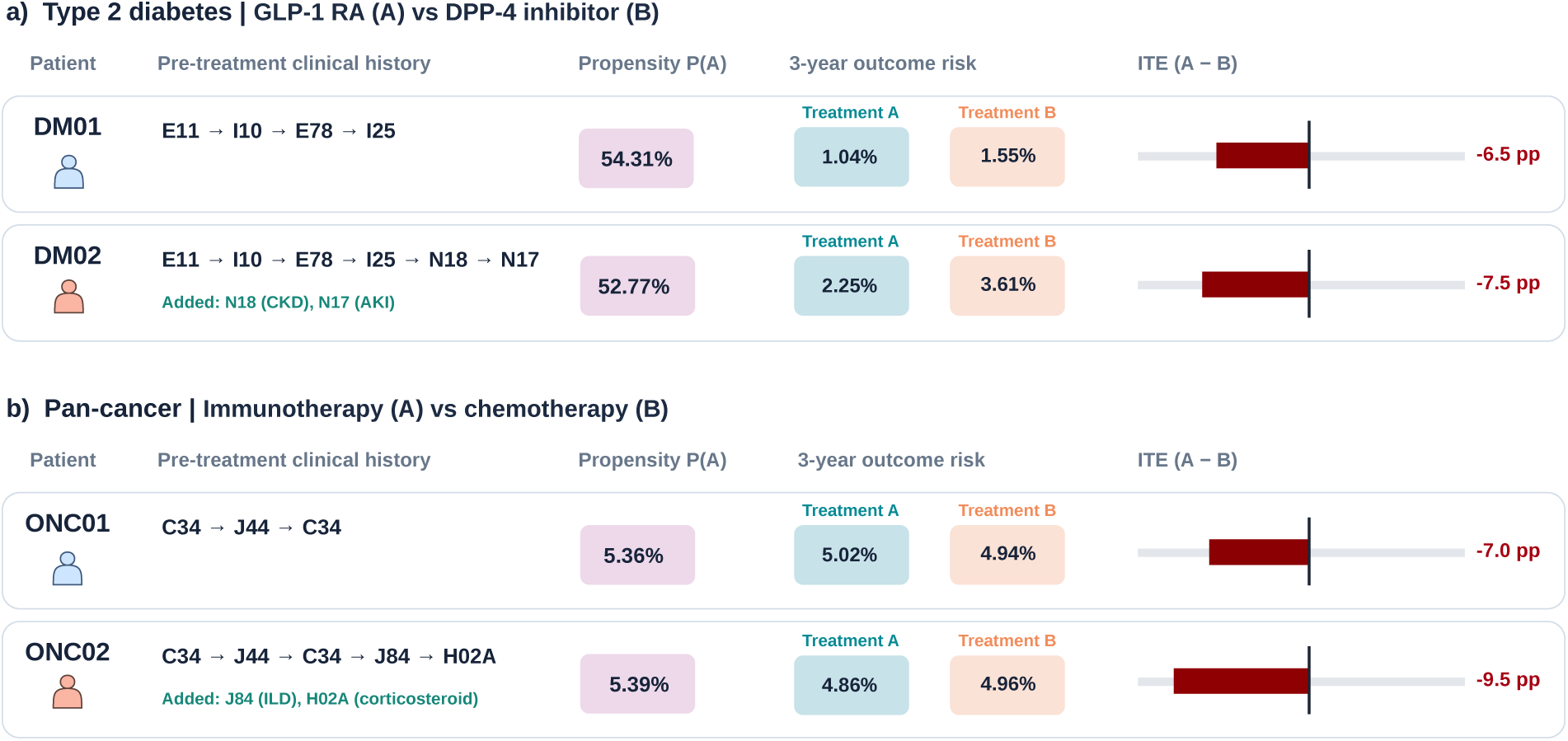
ITE predictions across different clinical histories. To demonstrate how longitudinal patient histories inform predictions of comparative treatment benefit, we consider example patient trajectories that differ in selected pre-treatment clinical events. **a**, Type 2 diabetes application comparing GLP-1 receptor agonists (treatment A) with DPP-4 inhibitors (treatment B). Two otherwise identical patient trajectories are compared, with the second additionally including chronic kidney disease (ICD-10 N18) and acute kidney injury (N17). The outcome is major adverse cardiovascular events (MACE) within three years after treatment initiation. **b**, Pan-cancer application comparing immunotherapy (treatment A) with chemotherapy (treatment B). Two patient trajectories with lung cancer are compared, with the second additionally including interstitial lung disease (ICD-10 J84) and prior systemic corticosteroid exposure (ATC H02A). The outcome is all-cause mortality within three years after treatment initiation. For each patient, the figure shows the predicted probability of receiving treatment A (propensity score), the predicted outcome risks under both treatment alternatives, and the ITE estimated by the second-stage CATE learner. Negative treatment effects indicate a lower predicted adverse outcome risk under treatment A, whereas positive values favor treatment B. Patient trajectories are synthetically constructed using temporally ordered clinical concepts, with all events occurring before treatment initiation. The patient trajectories were synthetically created to comply with the *All of Us Research Program*.

For the type 2 diabetes application, we considered two otherwise identical patient trajectories, where one additionally included diagnoses of chronic kidney disease and acute kidney injury (Figure 7a). For the pan-cancer application, we compared two patients with lung cancer, where one additionally had a history of interstitial lung disease and systemic corticosteroid exposure (Figure 7b). In the illustrative diabetes example, the additional renal complications were associated with a greater predicted benefit of GLP-1 receptor agonists, which is clinically plausible given the elevated cardiovascular risk associated with kidney disease. In the cancer example, the addition of interstitial lung disease and prior systemic corticosteroid exposure was associated with a larger predicted benefit of immunotherapy over chemotherapy (ITE *−*9.5 vs. *−*7.0 percentage points). This direction contrasts with prevailing clinical caution regarding immunotherapy in patients with pre-existing interstitial lung disease or corticosteroid exposure, a subgroup that has been largely excluded from pivotal immunotherapy trials. Conventional analyses that estimate average treatment effects across broad populations would not detect such patient-specific differences, and the ITE-Transformer may therefore reveal clinically relevant heterogeneity that remains masked by traditional methods. Importantly, this finding should be regarded as hypothesis-generating and requires external and prospective validation before it can inform treatment decisions.

These examples illustrate how the ITE-Transformer can be used to investigate whether additional clinical events are associated with changes in overall predicted outcome risks, the relative benefit of alternative treatments, or even the predicted direction of treatment benefit.

## Discussion

In this work, we introduced the ITE-Transformer, a transformer-based framework that combines self-supervised pre-training on longitudinal EHR trajectories with doubly robust causal machine learning to predict ITEs. We evaluated the ITE-Transformer across two clinically different settings (i.e., GLP-1 receptor agonists versus DPP-4 inhibitors in patients with type 2 diabetes, and immunotherapy versus chemotherapy in a pan-cancer cohort). Across both settings, the ITE-Transformer improved the discrimination and calibration of treatment assignment and outcome models, compared with conventional, tabular machine learning baselines and an otherwise identical transformer without pre-training. The ITE-Transformer identified substantial heterogeneity in predicted ITEs across patients. Together, these findings demonstrate the potential of integrating causal machine learning [2, 38, 39] with longitudinal EHR foundation models to support patient-level comparative effectiveness assessments and thus support real-world evidence generation. By identifying patients with different predicted treatment responses, ITE-Transformer can further generate hypotheses to guide prospective studies of personalized treatment strategies.

Existing clinical foundation models for EHR data have primarily focused on predicting future diagnoses [7–13, 40] or claims information [11], or predicting how likely patients respond to a given treatment [9, 14]. Although clinically informative, these approaches do not directly answer the counterfactual question of how patient outcomes would differ under different alternative strategies. Our ITE-Transformer addresses this gap by combining a pre-trained transformer with a doubly robust causal learning framework to predict ITEs from longitudinal patient histories.

Our results highlight the importance of self-supervised pre-training for ITE prediction. Compared with an otherwise identical transformer trained from scratch, the pre-trained ITE-Transformer led to more stable optimization and achieved a better performance in terms of discrimination and calibration metrics. This suggests that pre-training on large longitudinal EHR trajectories yields transferable representations of diagnoses, medication histories, comorbidities, and temporal disease patterns that remain informative when the downstream cohorts for treatment effect estimation are substantially smaller than the pre-training population. Our analyses of the learned embeddings further support this interpretation, where clinically related diagnoses and medication are mapped into similar, meaningful clusters, despite being learned only through self-supervised next-event prediction.

The improved discrimination and calibration of propensity and outcome models are important for the reliability of the doubly robust ITE estimation. In observational EHR data, treatment assignment is non-random and depends on clinical indications, disease severity, comorbidities, access patterns, and physician or patient preferences; accurately modeling treatment assignment and outcomes helps account for these systematic differences between treatment groups and reduce bias from measured confounding [16, 41]. Importantly, errors in propensity and outcome predictions can propagate through doubly robust pseudo-outcomes to the estimated treatment effects [20, 42], which explains the relevance of accurate and well-calibrated nuisance components. Our results suggest that ITE-Transformer captures clinically relevant information underlying treatment effect heterogeneity from longitudinal patient histories beyond conventional tabular approaches.

The ITE-Transformer predicted substantial heterogeneity in ITEs across both clinical settings. Some patients showed lower predicted MACE risk under GLP-1 receptor agonists than under DPP-4 inhibitors, while others showed smaller benefits or were predicted to benefit more from DPP-4 inhibitors. Similarly, in the pan-cancer cohort, the predicted mortality benefit of immunotherapy relative to chemotherapy varied across patients. These findings are consistent with the clinical expectation in both settings that the treatment response varies with a patient’s pre-treatment health status, comorbidities, prior medications, and disease histories [29, 34].

Our study has limitations, many of which are inherent to generating clinical evidence using real-world data [43]. First, although the pre-training cohort is large and diverse, it is drawn from a single data resource, which may limit the generalizability to other healthcare settings. Second, interpreting the predicted treatment effects causally requires assumptions that cannot be fully verified in observational data [2, 16, 17]. Although ITE-Transformer accounts for a broad set of demographic characteristics and longitudinal diagnosis and medication histories, unmeasured or incompletely recorded confounders may still bias treatment effect estimates. The results should therefore be interpreted as confounder-adjusted, model-based estimates for hypothesis generation rather than validated causal effects [44]. Clinical translation therefore requires rigorous validation of the predicted treatment effects using established approaches for real-world evidence generation, including sensitivity analyses (e.g., E-values [45]), target trial emulation [46, 47], and negative control outcomes [48]. Third, our model uses structured diagnoses and medication records; future research may include additional modalities such as laboratory measurements, procedures, clinical notes, imaging, or genomics. Fourth, validating ITE predictions is inherently challenging because the counterfactual outcome for each patient is unobserved. We therefore followed best practice [2, 23] for evaluating causal ML models by assessing propensity score overlap and effect score calibration [24], but external and prospective evaluation are left for future work. Our predictions should therefore primarily serve to generate hypotheses for future studies of personalized treatment strategies.

Overall, the ITE-Transformer is a clinical foundation model to predict ITEs from longitudinal EHR data, thereby advancing real-world evidence generation beyond conventional risk prediction. The ITE-Transformer provides a scalable framework for identifying who may benefit from alternative therapies to support hypothesis generation and personalized care.

## Methods

### Data

#### Patient cohort

We used data from the *All of Us Research Program* [19], with all analyses conducted within the *All of Us Researcher Workbench* [49]. At the time of study, the program included 626,383 participants aged 18 years or older recruited from a more than 340 sites. The available data are de-identified and include participant surveys, physical measurements, and electronic health records (EHRs). For this study, we included the 350,000 participants with available structured EHR data. Cohort characteristics are reported in the Supplementary Table S1 and Supplementary Figure S1. Data preprocessing complied with all relevant regulation of the *All of Us Research Program*. No ethical approval was necessary at the host institution due to the de-identified, non-interventional design of the study.

#### Data preprocessing

We represented longitudinal patient histories using diagnosis and medication codes aggregated at clinically interpretable levels. Diagnoses were encoded using 1,257 level-3 ICD codes (e.g., E11 – “Type 2 diabetes mellitus”) from chapters I–XVII. Medication exposures were represented using 215 level-3 ATC codes (e.g., A10A – “Insulins and analogues”) across all ATC chapters. Together, these codes formed a vocabulary of 1,472 clinical concepts, which was used to balance clinical specificity with reduced sparsity. Clinical concepts were mapped to the respective coding hierarchies using standardized OMOP vocabulary tables available in the *All of Us Researcher Workbench*. The included ICD and ATC chapters are listed in Supplementary Tables S2 and S3.

To reduce redundant clinical events, we applied temporal deduplication within each patient trajectory (similar to but less strict as in Delphi-2M [10]). For diagnoses, we retained at most one occurrence of each code within a six-month window. For medications, we retained only the first recorded exposure to each medication concept.

#### Study cohorts

We evaluated the ITE-Transformer in two comparative-effectiveness tasks designed as target trial emulations [46]. Each analysis was defined by a target patient population, a study treatment, an active comparator, and a clinical outcome. Following an active-comparator, new-user design [17], we defined the treatment index date as initiation of either treatment and used only pre-treatment clinical histories as model input [46]. Eligible patients were required to have the relevant diagnosis before treatment initiation and at least one recorded exposure to either the study treatment or comparator. Patients who initiated both treatment classes on the same date or had otherwise ambiguous treatment assignment were excluded.

We considered the following two treatment comparisons:

- **Type 2 diabetes cohort.** GLP-1 receptor agonists and DPP-4 inhibitors are used at comparable stages in the treatment pathway for type 2 diabetes [50], yet differ in their cardiovascular benefits. GLP-1 receptor agonists have been shown to reduce major adverse cardiovascular events (MACE), while DPP-4 inhibitors showed no corresponding benefit [28, 51], but studies reported heterogeneity in treatment response [29]. Patients diagnosed with type 2 diabetes mellitus (ICD-10 code E11) initiating either a GLP-1 receptor agonist or a DPP-4 inhibitor were included, with the treatment index date defined as the first recorded prescription of either drug class following the diabetes diagnosis. The outcome was the occurrence of a major adverse cardiovascular event (MACE) within three years after treatment initiation.
- **Pan-cancer cohort.** Patients diagnosed with a solid malignant neoplasm were identified using the corresponding ICD-10 codes. Eligible patients initiated either immunotherapy or chemotherapy, with the treatment index date defined as the first recorded administration of either treatment following cancer diagnosis. The outcome was all-cause mortality within three years after treatment initiation.

Details are provided in the supplements regarding eligibility criteria (Supplementary Tables S4 and S5), treatment definitions (Supplementary Tables S6 and S7), and outcome definitions (Supplementary Table S8).

### ITE-Transformer

#### Overview

The ITE-Transformer consists of four main components. First, longitudinal EHRs are represented as temporally ordered sequences of diagnoses and medication exposures, together with information on patient age and sex (*patient trajectory representation*). Second, these trajectories are processed by a decoder-only transformer that produces contextualized representations of clinical events and patient histories (*transformer architecture*). Third, the transformer is pre-trained on a large EHR cohort using self-supervised next-event prediction to learn general representations of clinical concepts in longitudinal patient trajectories (*self-supervised pre-training*). Fourth, the pretrained transformer is leveraged for comparative effectiveness assessments using a doubly robust causal learning framework (*treatment effect prediction*). In the fourth component, the transformer-derived patient representations are first used to predict treatment propensity and treatment-specific outcome risks, which are subsequently combined into doubly robust pseudo-outcomes to train a model for predicting ITEs (defined by the conditional average treatment effect; CATE).

These first three components are adapted from recent clinical foundation models [9–11]. The key methodological novelty of the ITE-Transformer is the fourth component, which allows ITE-Transformer not only to use patient trajectories for risk or outcome modeling but for treatment effect modeling. As a result, we integrate principles from causal machine learning [2,18] to predict ITEs under different treatment strategies. This enables ITE-Transformer to make individualized assessments of the comparative effectiveness of treatments from longitudinal patient trajectories.

#### Patient trajectory representation

Following recent longitudinal EHR foundation models [9–11], we represent each patient as a chronologically ordered sequence of clinical events. Each event corresponds to a diagnosis or medication token from a predefined clinical vocabulary and is paired with the patient’s age at the time of the event. A token encoding the biological sex is added at the beginning of the sequence.

Clinical tokens are mapped to learnable vector embeddings and combined with a continuous age-based encoding [10]. Unlike conventional positional encodings, which capture the order of events, the age-based encoding also captures the relative timing and therefore accommodates for the irregular timing between clinical events.

#### Transformer architecture

Patient trajectories are encoded using a decoder-only transformer with causal self-attention [52]. The architecture consists of stacked transformer blocks with multi-head self-attention, position-wise feed-forward layers, residual connections, and layer normalization. Causal masking restricts each position to information observed at or before that time point in the trajectory, thus preventing that information from future events from entering the representation.

The transformer produces a contextualized hidden representation for each token in the patient trajectory. During self-supervised pre-training, these representations are used to predict subsequent clinical events. For the downstream task of predicting treatment effects, we use the hidden state of the last non-padded token before treatment initiation as a fixed-dimensional representation of the patient’s pre-treatment clinical history.

A schematic overview of the architecture is provided in Supplementary Figure S2. Hyper-parameters, including the number of transformer layers, attention heads, embedding dimension, feed-forward dimension, dropout, and total number of trainable parameters, are reported in Supplementary Table S9.

#### Self-supervised pre-training

The transformer is pre-trained on longitudinal EHR trajectories before being adapted to treatment effect estimation. We use autoregressive next-event prediction as the self-supervised learning objective. At each position in a patient trajectory, the transformer receives the clinical history observed up to that point and predicts the next diagnosis or medication token. This objective encourages the transformer to learn general temporal and clinical structure from patient histories, including relationships among diagnoses, medication exposures, demographics, and disease timing, without requiring task-specific outcome labels.

Formally, let *x*_1_ denote the sequence of clinical tokens for patient *i*, and let *p_θ_*(*x_t_*_+1_ *| x*_1_) denote the probability of the subsequent clinical token given the preceding sequence. The pre-training objective minimizes the average negative log-likelihood over valid sequence positions, i.e.,

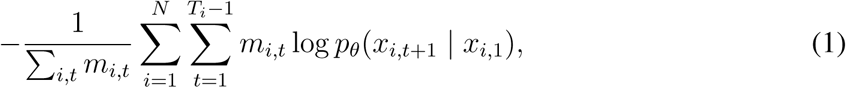

where *m_i,t_* indicates whether the corresponding target token is non-padded. Padding positions are excluded from the loss.

#### ITE prediction

##### Estimand

The ITE-Transformer leverages the pre-trained longitudinal patient representations to predict ITEs. Let *X* denote the transformer-derived representation of a patient’s pre-treatment clinical history, *A ∈* 0, 1 the observed treatment assignment (i.e., *A* = 1 denotes the study treatment and *A* = 0 the comparator), and *Y ∈* 0, 1 the observed clinical outcome (i.e., *Y* = 1 indicates occurrence of the task-specific endpoint during follow-up). Formally, we aim to learn the conditional average treatment effect (CATE)

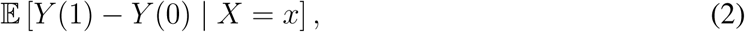

where *Y* (1) and *Y* (0) denote the potential outcomes under the study treatment and comparator, respectively. The CATE *τ* (*x*) then represents the expected difference in outcomes between the two treatment strategies for patients with a given pre-treatment representation. Throughout this paper, we use the term ITE to refer to a patient-specific prediction of the conditional average treatment effect; note that this is different from the unobservable individual treatment effect.

##### Doubly robust learner

We estimate the CATE using a doubly robust (DR) learner based on augmented inverse probability weighting (AIPW) [18, 20, 21]. The DR-learner consists of two stages. In the first stage, the *propensity model*

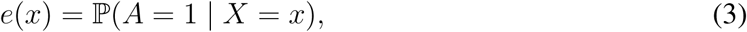

estimates the probability of receiving the study treatment conditional on the observed clinical history, and the *treatment-specific outcome models*

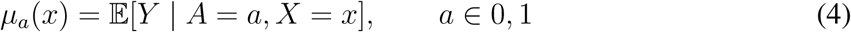

estimate the expected outcome under each treatment alternative. The estimated propensity and outcome functions, *e*^(*x*), *µ*^_1_(*x*), and *µ*^_0_(*x*), are together referred to as nuisance components, which are required for treatment effect estimation but are not themselves the primary quantities of interest.

In the second stage, these predictions *e*^(*x*), *µ*^_1_(*x*), and *µ*^_0_(*x*) are used to construct the *doubly robust pseudo-outcome*

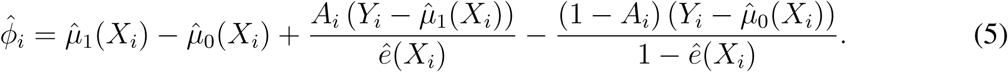

The first term captures the difference between the treatment-specific outcome predictions, while the inverse-propensity-weighted residuals use the observed treatment and outcome to correct for errors in the outcome models. In the second stage, a CATE learner is trained on these pseudo-outcomes to predict how ITEs vary across patient histories. Under the causal assumptions of consistency, conditional exchangeability, and positivity, the estimator is doubly robust; i.e., it remains consistent if either the propensity model or the treatment-specific outcome models are consistently estimated [17, 20].

##### Task-specific adaptation

For each comparative-effectiveness task, the pre-trained ITE-Transformer is adapted to the corresponding treatment cohort using the above two-staged doubly robust approach.

To reduce overfitting bias and enable valid inference, we use *K*-fold cross-fitting [53] when estimating the propensity and outcome models and constructing the pseudo-outcomes. For this, the development cohort is partitioned into *K* folds; for each fold, the propensity and outcome models are trained on the remaining *K −* 1 folds and used to generate predictions for the held-out fold. Each patient’s pseudo-outcome is therefore constructed from propensity and outcome predictions generated by a model that was not trained on that patient.

For the first stage, we attach a task-specific prediction head with three output logits to the transformer in order to produce estimates *e*^(*X*), *µ*^_1_(*X*), and *µ*^_0_(*X*). The propensity model is trained using binary cross-entropy for the observed treatment assignment, while the outcome loss is computed only for the treatment actually received. The joint training objective is

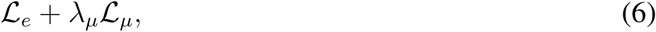

where *L_e_*denotes the propensity loss, *L_µ_*the factual outcome loss, and *λ_µ_*controls their relative contribution.

In the second stage, a CATE prediction head is trained on the cross-fitted pseudo-outcomes using the transformer-derived patient representation as input. The transformer backbone is frozen during this stage, and the CATE prediction head is optimized using the Huber loss. The resulting model maps the pre-treatment patient representation to an ITE prediction.

### Baselines

We compared the ITE-Transformer with four baselines using identical data splits and cross-fitting procedure. The comparisons focused on the propensity and outcome predictions, which can be evaluated directly against observed data by comparing model discrimination and calibration. This allows us to assess whether ITE-Transformer provides more reliable estimates of the nuisance components used for subsequent treatment effect estimation tasks, without the need to have information about unobserved counterfactual outcomes.

1. *Age and sex classifier*. Following prior work on clinical foundation models [9, 10], we trained classifiers using only patient age at treatment initiation and sex to predict treatment assignment and observed outcomes as a minimal baseline.
2. *Random forest*. We trained a random forest classifier on fixed-width tabular representations of the pre-treatment history, consisting of binary indicators for the most prevalent clinical codes together with summary features of the patient trajectory. Separate models were trained for propensity and outcome prediction. The models were implemented using scikit-learn [54]. Hyperparameters are reported in Supplementary Table S13.
3. *XGBoost*. We trained an XGBoost classifier on the same tabular representation as the random forest baseline to provide a strong nonlinear benchmark [55]. Separate models were again trained using the XGBoost implementation from [56] for propensity and outcome prediction. Hyperparameters are reported in Supplementary Table S14.
4. *Non-pretrained transformer (NP-Transformer).* To isolate the contribution of self-supervised pre-training, we trained an otherwise identical transformer with the same architecture, vocabulary, age encoding, and prediction heads as the ITE-Transformer, but with randomly initialized weights and no self-supervised pre-training. This baseline was trained directly on each downstream cohort using the same supervised first-stage objective. Hyperparameters are reported in Supplementary Table S12.

### Evaluation

Following established practice for causal machine learning [2], we evaluated the ITE-Transformer as follows. First, we assessed the predictive performance of the propensity model and treatment-specific outcome models on the held-out folds. Discrimination was quantified using the area under the receiver operating characteristic curve (AUROC) and the area under the precision–recall curve (AUPRC) [57]. Calibration was assessed by grouping patients according to their predicted probabilities and comparing the mean predicted probability within each group with the corresponding observed treatment or outcome frequency [58]. We additionally examined the distributions of estimated propensity scores across treatment groups to assess empirical overlap [59].

Second, we assesed the effect score calibration [24] to examine whether predicted ITEs aligned with stratum-specific average treatment effects across patient strata with varying predicted treatment benefit. Intuitively, well-calibrated effect scores should align with the predicting treatment effect within each patient stratum. To assess this, patients were ranked by their predicted ITEs and divided into strata. Within each stratum, the mean over the predicted ITEs was compared with the corresponding average treatment effect (ATE) estimated using augmented inverse probability weighting (AIPW) [42]. This analysis evaluates whether the CATE learner captures systematic variation in treatment effects across patient strata.

Experiments were repeated across five random seeds, using the same seeds across models to allow consistent comparisons.

### Interpretability of the predicted ITEs

To examine which clinical factors of the pre-treatment clinical history contributed to the predicted ITEs, we used SHapley Additive exPlanations (SHAP) [26]. For this, we aggregated diagnosis and medication concepts to higher-level ICD and ATC groups to reduce sparsity and thus improve interpretability. SHAP values were then used to quantify the contribution of each feature group to the predicted ITE for individual patients. The magnitude of a SHAP value reflects the strength of a feature’s contribution, while the corresponding sign indicates whether the feature shifts the prediction toward a larger relative benefit from the study treatment or from the comparator.

### Implementation details

All analyses were performed in Python. We implemented the ITE-Transformer using PyTorch [60]. The model is optimized using AdamW [61], gradient clipping with a maximum norm of 1.0, and a cosine learning-rate schedule with linear warm-up. For computational efficiency, trajectories are represented using a fixed maximum context length. For sequences exceeding this limit, we retain the most recent clinical history; shorter trajectories are padded to the maximum sequence length. Padding tokens are excluded from both attention and loss computation. Hyperparameters were selected through empirical tuning to balance validation performance and computational efficiency; see Supplementary Table S9.

For the task-specific adaptations, separate learning rates were used for the pre-trained transformer backbone and the newly initialized prediction heads to preserve learned representations while adapting to each treatment comparison. Estimated propensity scores were clipped [62] before construction of the AIPW pseudo-outcomes to reduce instability from extreme inverse probability weights. Further training details, including learning rates, batch sizes, number of epochs, dropout, and weight decay, are reported in Supplementary Tables S10 and S11.

## Data availability

The data used in this study were obtained from the *All of Us Research Program* [19] ([Registered] Tier Dataset v8) and are available to authorized researchers through the *All of Us Researcher Work-bench* (https://workbench.researchallofus.org/), subject to the program’s data access requirements. Individual-level patient data cannot be publicly shared due to privacy and data use restrictions.

## Code availability

The code developed for this study, including the implementation of ITE-Transformer, baseline methods, and evaluation procedures, is publicly available in a GitHub repository at https://github.com/sfeuerriegel/ITE-Transformer.

## Acknowledgments

We gratefully acknowledge the participants of the All of Us Research Program for their contributions to this research. We also thank the National Institutes of Health’s All of Us Research Program for providing access to the data used in this study.

## Author contributions

S.F. had the idea. C.A. implemented the model and performed the analysis. All authors contributed to conceptualization, manuscript writing, and approved the manuscript.

## Competing interests

The authors declare no competing interests.

## Supplementary Materials

### Pre-training cohort

**Table S1:**
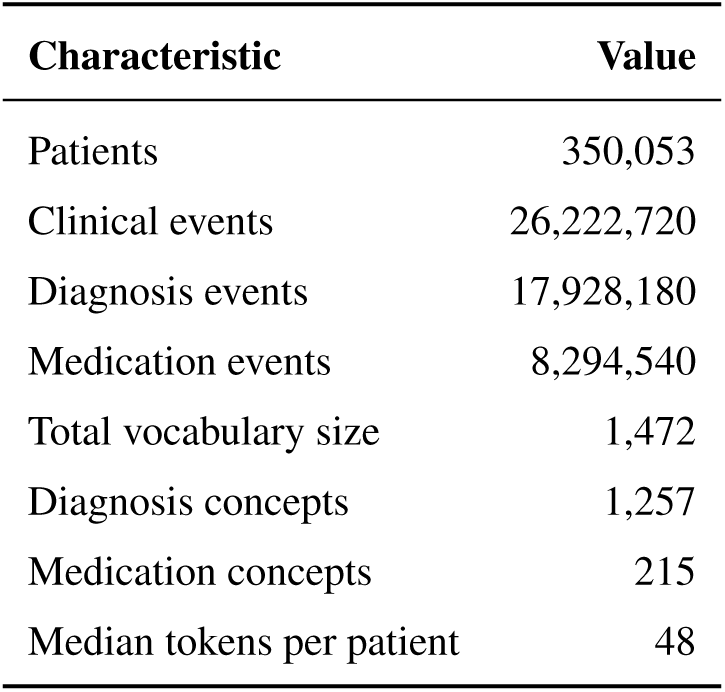
Summary statistics of the pre-training cohort and model vocabulary.

**Figure S1:**
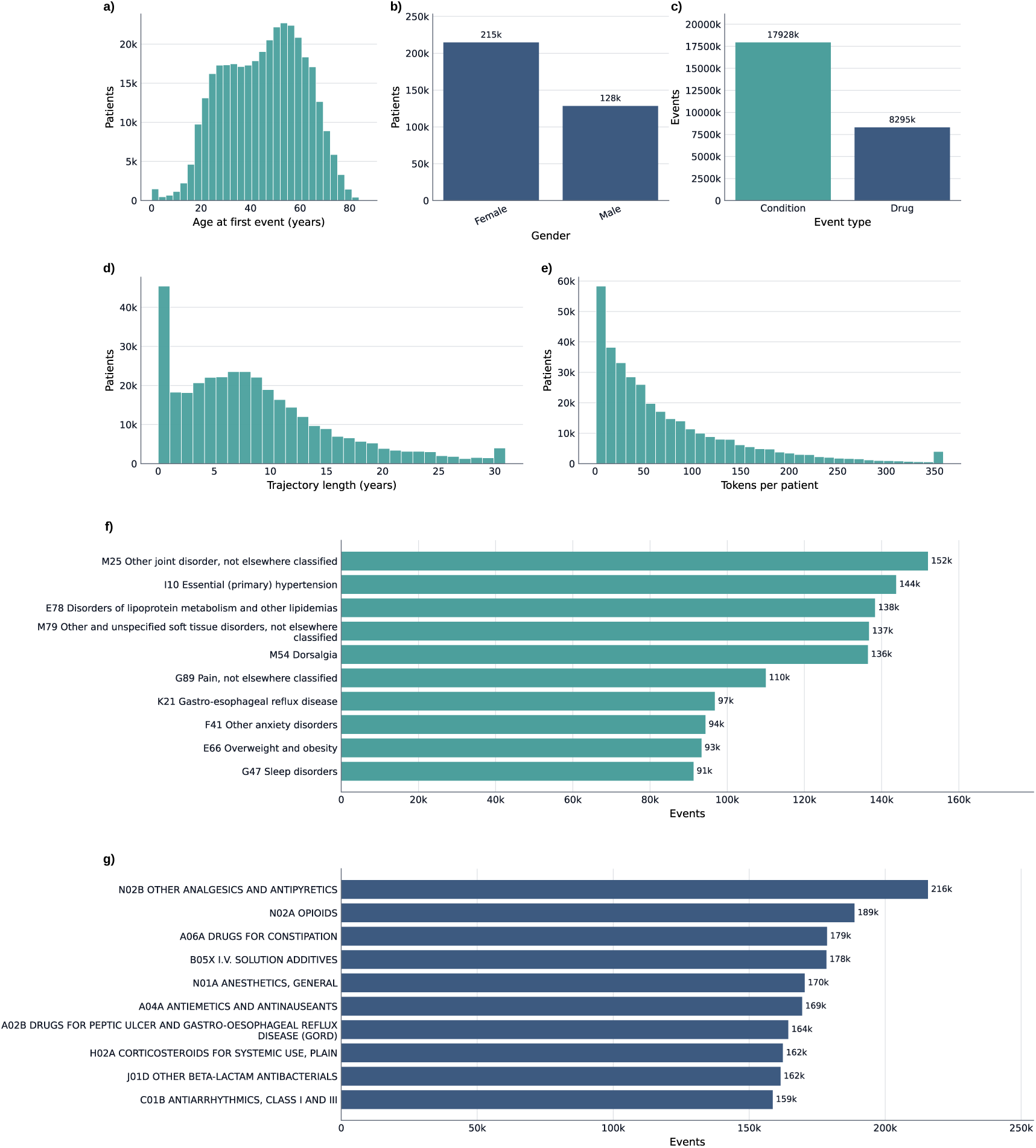
Characteristics of the pre-training cohort. **a**, Distribution of age at first recorded clinical event. **b**, Sex distribution. **c**, Length of patient trajectories in years, measured from the first to the last recorded event. **d**, Distribution of number of tokens per patient. **e**, Distribution of clinical event types. **f**, Top-10 most frequent diagnosis codes. **g**, Top ten most frequent medication exposure codes. Code frequencies in **f** and **g** are counted once per patient. For visualization, distributions are capped at the 99th percentile.

### Model architecture

**Figure S2:**
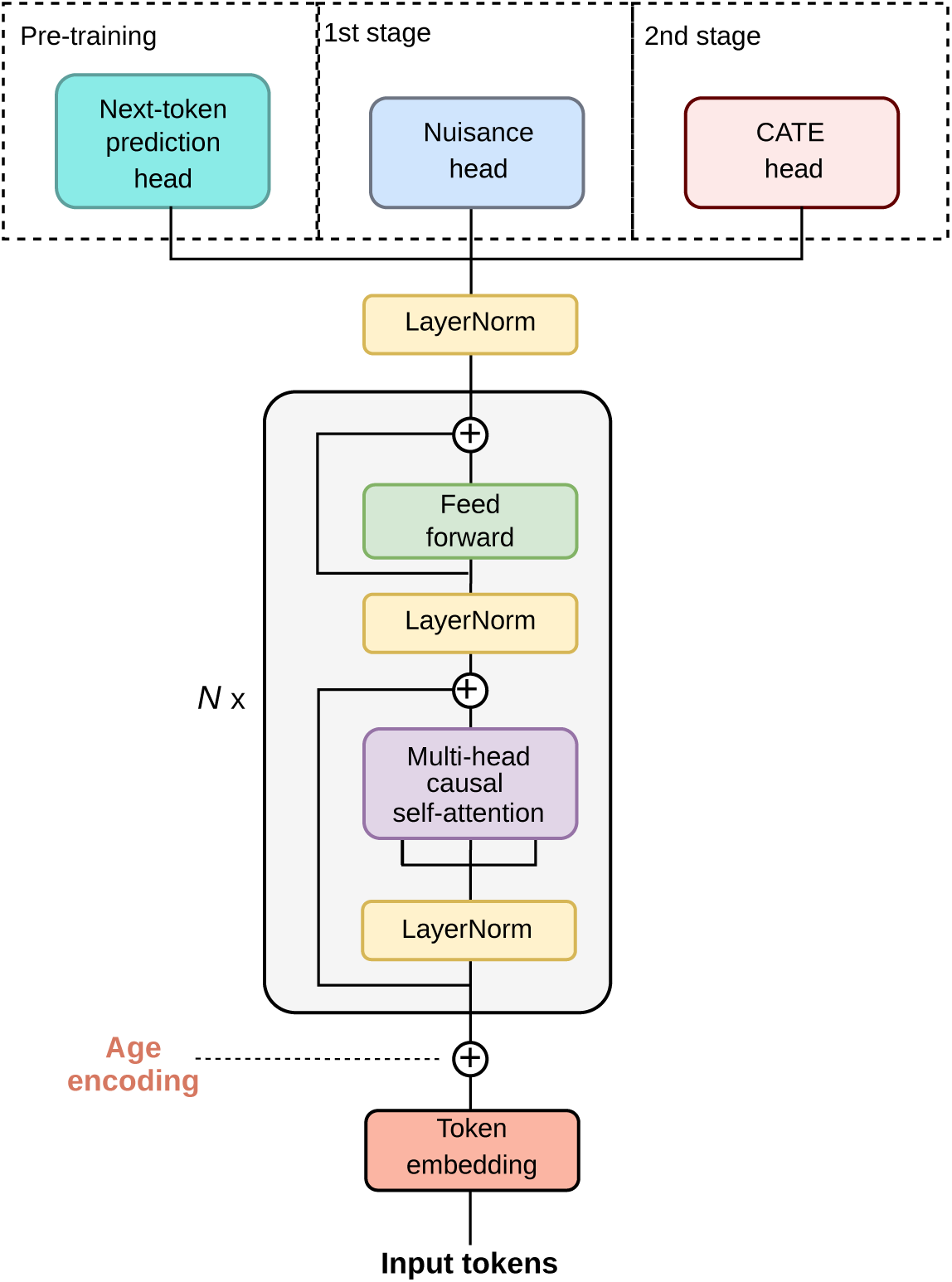
Overview of the ITE-Transformer architecture. Patient trajectories are represented as sequences of clinical tokens, combined with continuous age encodings and processed by a decoder-only transformer consisting of *N* stacked causal self-attention blocks. The resulting hidden representations are used for three purposes. (i) During pre-training, a linear output head projects the hidden representation to the vocabulary size for autoregressive next-token prediction. (ii) In the first stage of treatment effect estimation, a two-layer prediction head jointly predicts three binary quantities: the propensity for treatment assignment and the potential outcome probabilities under the study treatment and comparator treatment (i.e., the nuisance components). These predictions are used to construct doubly robust pseudo-outcomes. (iii) In the second stage, a two-layer CATE head is trained on the pseudo-outcomes to predict individualized treatment effects.

### ICD diagnosis chapters

**Table S2:** Overview of ICD diagnosis chapters I–XVII used for aggregating diagnosis codes.

| **Chapter** | **Identifying letter(s)** | **Chapter name** |
|---|---|---|
| I | A–B | Certain infectious and parasitic diseases |
| II | C–D | Neoplasms |
| III | D | Diseases of the blood and blood-forming organs and certain disorders involving the immune mechanism |
| IV | E | Endocrine, nutritional and metabolic diseases |
| V | F | Mental, behavioral and neurodevelopmental disorders |
| VI | G | Diseases of the nervous system |
| VII | H | Diseases of the eye and adnexa |
| VIII | H | Diseases of the ear and mastoid process |
| IX | I | Diseases of the circulatory system |
| X | J | Diseases of the respiratory system |
| XI | K | Diseases of the digestive system |
| XII | L | Diseases of the skin and subcutaneous tissue |
| XIII | M | Diseases of the musculoskeletal system and connective tissue |
| XIV | N | Diseases of the genitourinary system |
| XV | O | Pregnancy, childbirth and the puerperium |
| XVI | P | Certain conditions originating in the perinatal period |
| XVII | Q | Congenital malformations, deformations and chromosomal abnormalities |

### ATC medication chapters

**Table S3:**
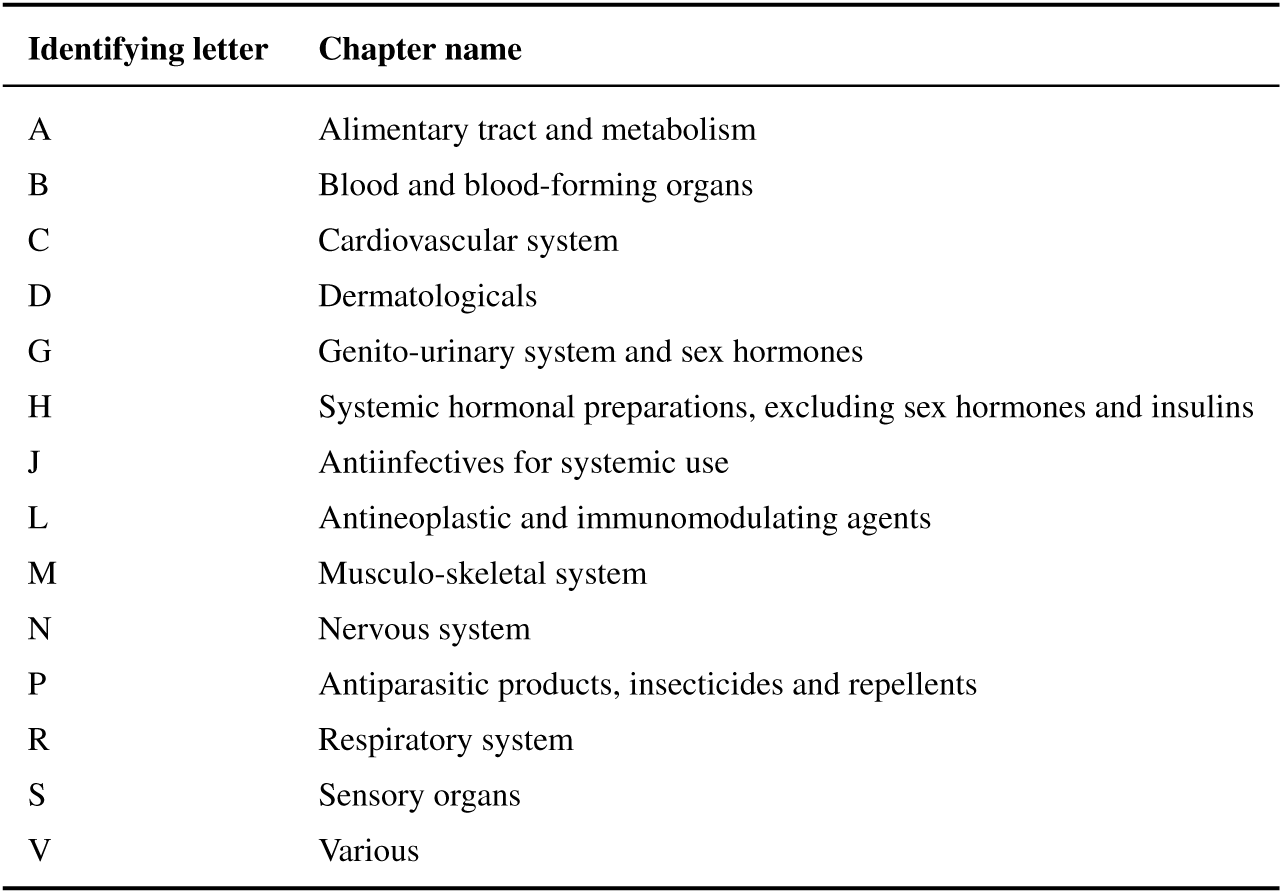
Overview of ATC medication chapters used for aggregating medication codes.

| **Identifying letter** | **Chapter name** |
|---|---|
| A | Alimentary tract and metabolism |
| B | Blood and blood-forming organs |
| C | Cardiovascular system |
| D | Dermatologicals |
| G | Genito-urinary system and sex hormones |
| H | Systemic hormonal preparations, excluding sex hormones and insulins |
| J | Antiinfectives for systemic use |
| L | Antineoplastic and immunomodulating agents |
| M | Musculo-skeletal system |
| N | Nervous system |
| P | Antiparasitic products, insecticides and repellents |
| R | Respiratory system |
| S | Sensory organs |
| V | Various |

### Cohort definitions for the clinical applications

**Table S4:** ICD diagnosis codes used to define the type 2 diabetes cohort for the GLP-1 receptor agonist versus DPP-4 inhibitor treatment comparison.

| **ICD code** | **Diagnosis name** |
|---|---|
| E11 | Type 2 diabetes mellitus |

**Table S5:** ICD diagnosis codes used to define the pan-cancer cohort for the immunotherapy versus chemotherapy treatment comparison.

| **ICD code** | **Diagnosis name** |
|---|---|
| C15 | Malignant neoplasm of esophagus |
| C16 | Malignant neoplasm of stomach |
| C17 | Malignant neoplasm of small intestine |
| C18 | Malignant neoplasm of colon |
| C19 | Malignant neoplasm of rectosigmoid junction |
| C20 | Malignant neoplasm of rectum |
| C21 | Malignant neoplasm of anus and anal canal |
| C22 | Malignant neoplasm of liver and intrahepatic bile ducts |
| C23 | Malignant neoplasm of gallbladder |
| C24 | Malignant neoplasm of other and unspecified parts of biliary tract |
| C25 | Malignant neoplasm of pancreas |
| C26 | Malignant neoplasm of other and ill-defined digestive organs |
| C30 | Malignant neoplasm of nasal cavity and middle ear |
| C31 | Malignant neoplasm of accessory sinuses |
| C32 | Malignant neoplasm of larynx |
| C33 | Malignant neoplasm of trachea |
| C34 | Malignant neoplasm of bronchus and lung |
| C35 | Malignant neoplasm of thymus |
| C36 | Malignant neoplasm of heart, mediastinum and pleura |
| C37 | Malignant neoplasm of other and ill-defined sites in the respiratory system and intrathoracic organs |
| C38 | Malignant neoplasm of bone and articular cartilage |
| C39 | Malignant neoplasm of mesothelial and soft tissue |
| C43 | Malignant melanoma of skin |
| C50 | Malignant neoplasm of breast |
| C51 | Malignant neoplasm of vulva |
| C52 | Malignant neoplasm of vagina |
| C53 | Malignant neoplasm of cervix uteri |
| C54 | Malignant neoplasm of corpus uteri |
| C55 | Malignant neoplasm of uterus, part unspecified |
| C56 | Malignant neoplasm of ovary |
| C57 | Malignant neoplasm of other and unspecified female genital organs |
| C58 | Malignant neoplasm of placenta |
| C60 | Malignant neoplasm of penis |
| C61 | Malignant neoplasm of prostate |
| C62 | Malignant neoplasm of testis |
| C63 | Malignant neoplasm of other and unspecified male genital organs |
| C64 | Malignant neoplasm of kidney, except renal pelvis |
| C65 | Malignant neoplasm of renal pelvis |
| C66 | Malignant neoplasm of ureter |
| C67 | Malignant neoplasm of bladder |
| C68 | Malignant neoplasm of other and unspecified urinary organs |

**Table S6:** Medications used to define GLP-1 receptor agonists and DPP-4 inhibitors exposure groups in the type 2 diabetes cohort.

| **Treatment class** | **ATC code** |
|---|---|
| GLP-1 receptor agonists | A10BJ |
| DPP-4 inhibitors | A10BH |

**Table S7:**
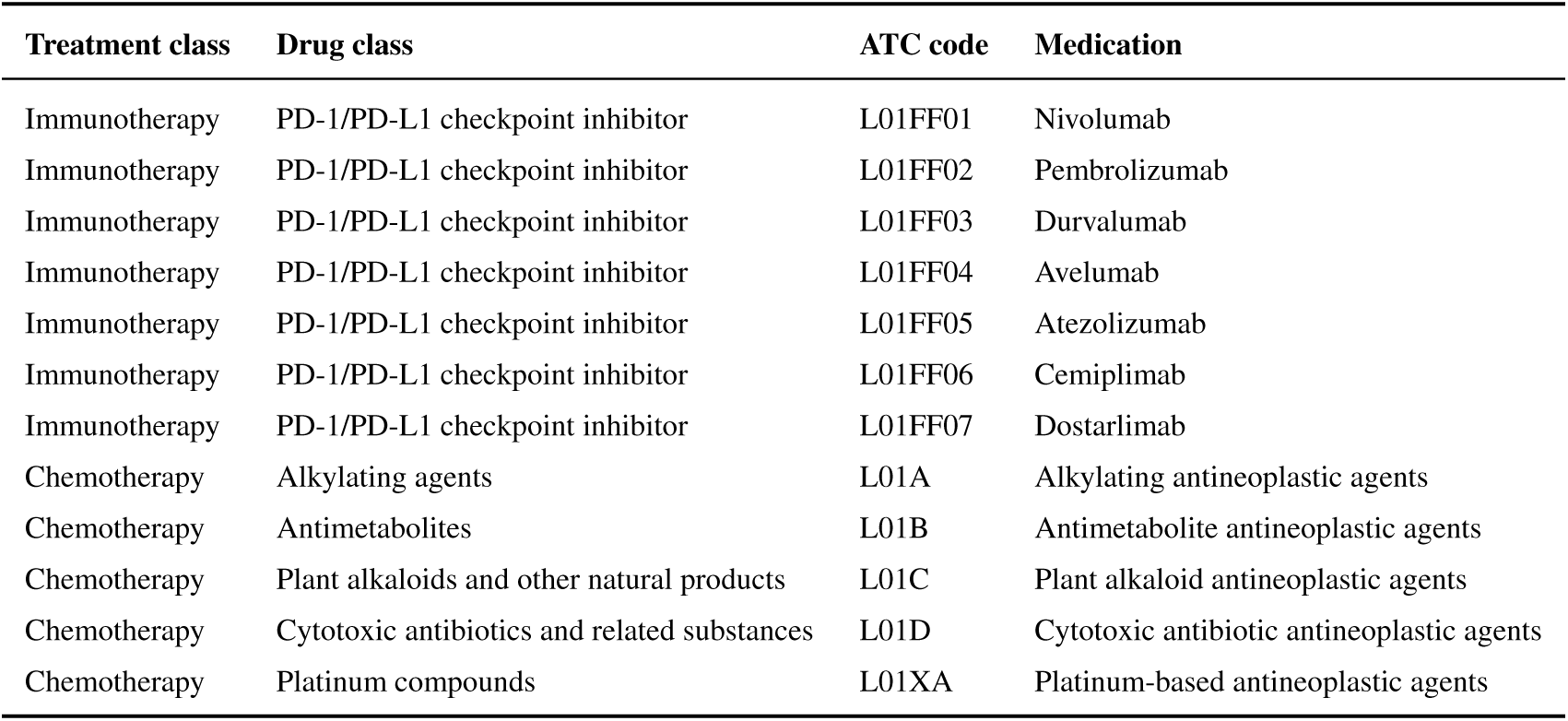
Medication concepts used to define immunotherapy and chemotherapy exposure groups in the pan-cancer cohort.

**Table S8:** Diagnoses codes used to define outcomes in the GLP-1 receptor agonists and DPP-4 inhibitors task.

| **ICD code** | **Diagnosis name** |
|---|---|
| I21 | Acute Myocardial Infarction |
| I22 | Subsequent ST elevation (STEMI) and non-ST elevation (NSTEMI) myocardial infarction |
| I23 | Certain current complications following ST elevation (STEMI) and non-ST elevation(NSTEMI) myocardial infarction (within the 28 day period) |
| I50 | Heart failure |

### Comparison of learning curves

**Figure S3:**
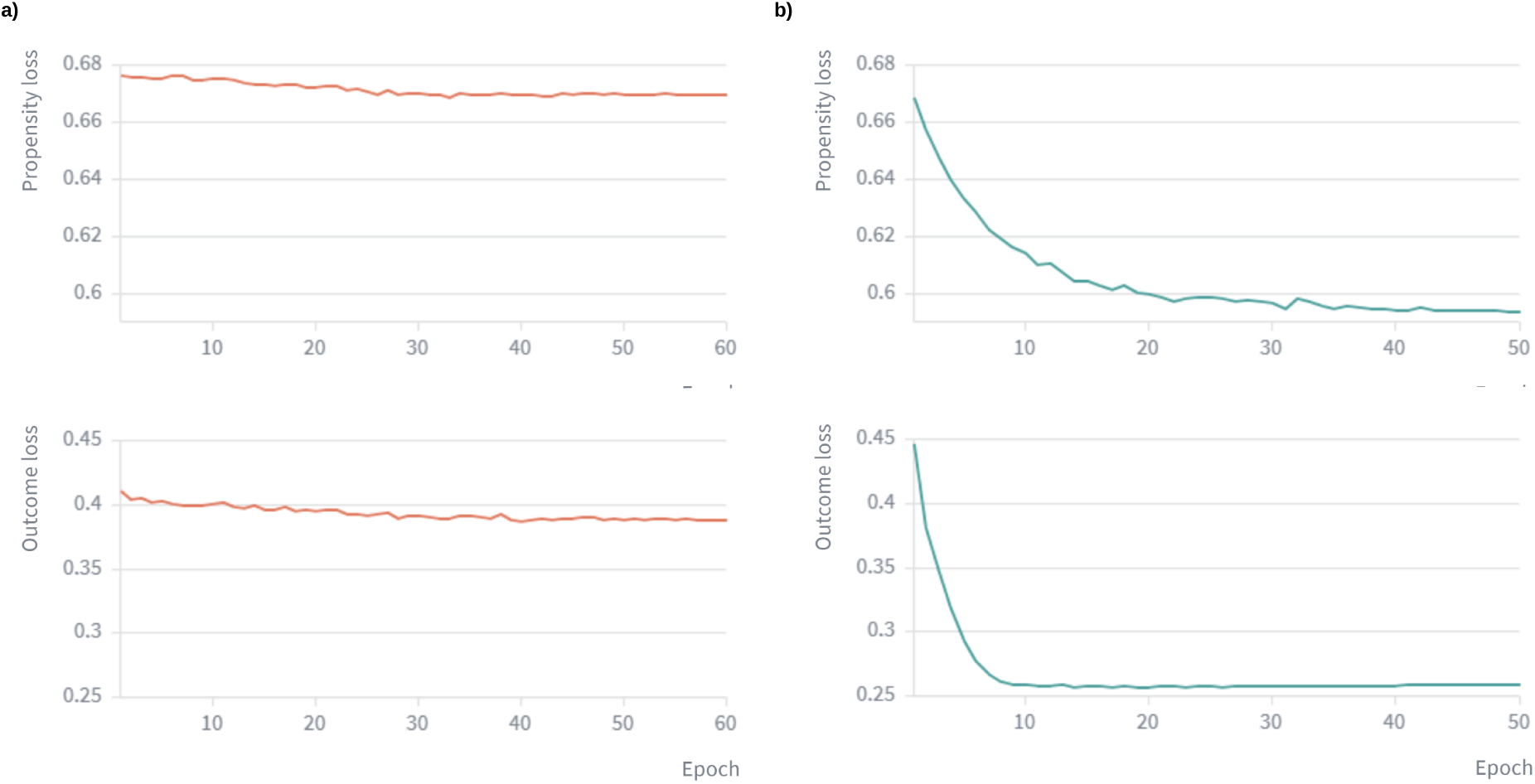
Representative validation loss curves with and without self-supervised pre-training. Validation losses for propensity and outcome models are shown across training epochs for a representative cross-validation fold (fold 0, random seed 17). **a**, Transformer trained without self-supervised pre-training (NP-Transformer). **b**, Pretrained ITE-Transformer. The example demonstrates that pre-training leads to faster convergence and lower validation losses.

### Empirical analysis of overlap

**Figure S4:**
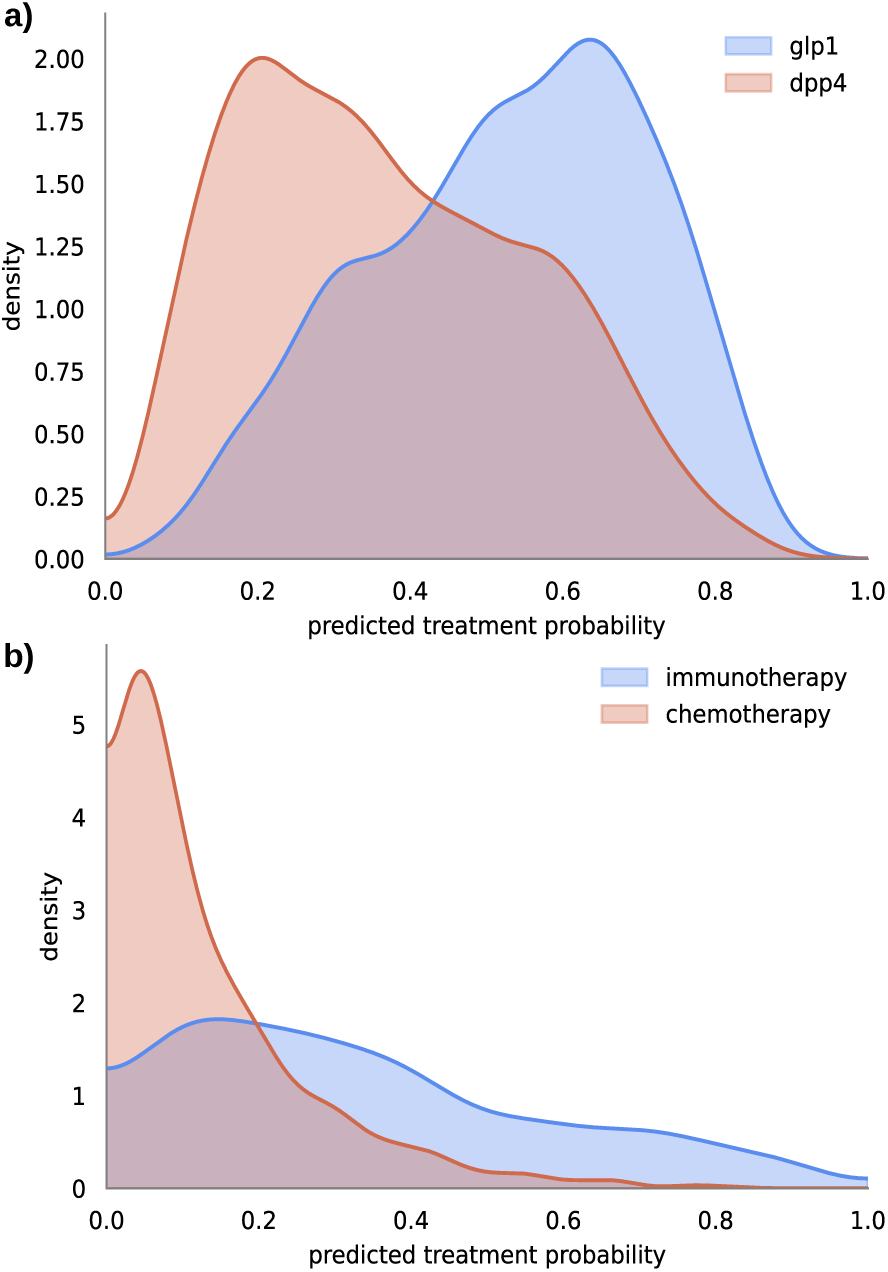
Predicted propensity scores by treatment assignment. To empirically assess whether there is sufficient overlap between treatment groups, we examined the distributions of predicted propensity scores. Propensity scores represent the estimated probability of receiving the study treatment conditional on the observed pre-treatment patient history. We find sufficient overlap, which supports the positivity assumption. **a**, GLP-1 receptor agonists versus DPP-4 inhibitors task. **b**, Immunotherapy versus chemotherapy task.

### Pseudo-outcomes

**Figure S5:**
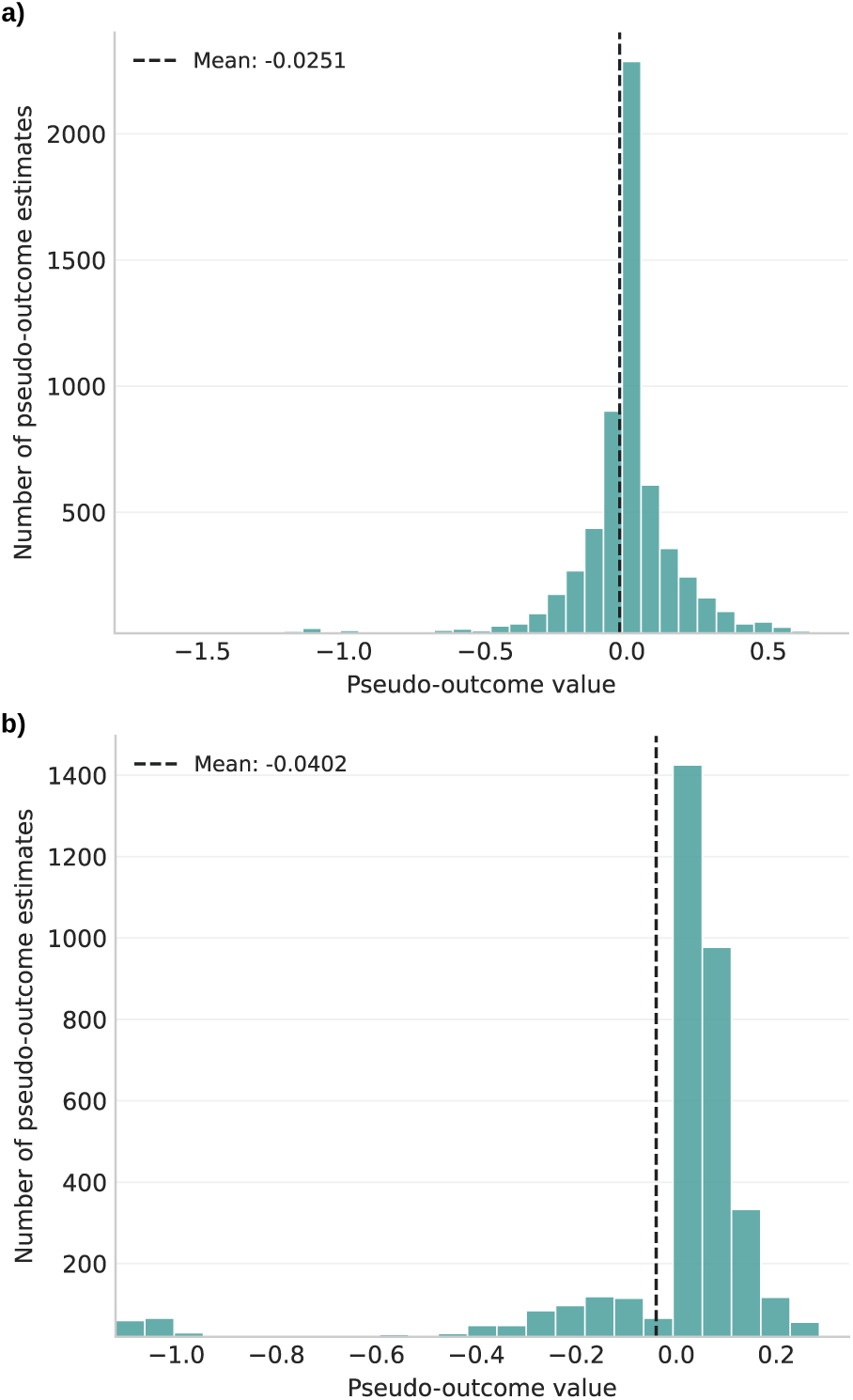
Distribution of pseudo-outcomes. **a**, GLP-1 receptor agonists vs DPP-4 inhibitors task. **b**, Immunotherapy vs chemotherapy task. To comply with dissemination regulations of the *All of Us Research Program*, the y-axis in histograms are capped at 20 patients.

### Comparison of predicted ITEs with causal forests

**Figure S6:**
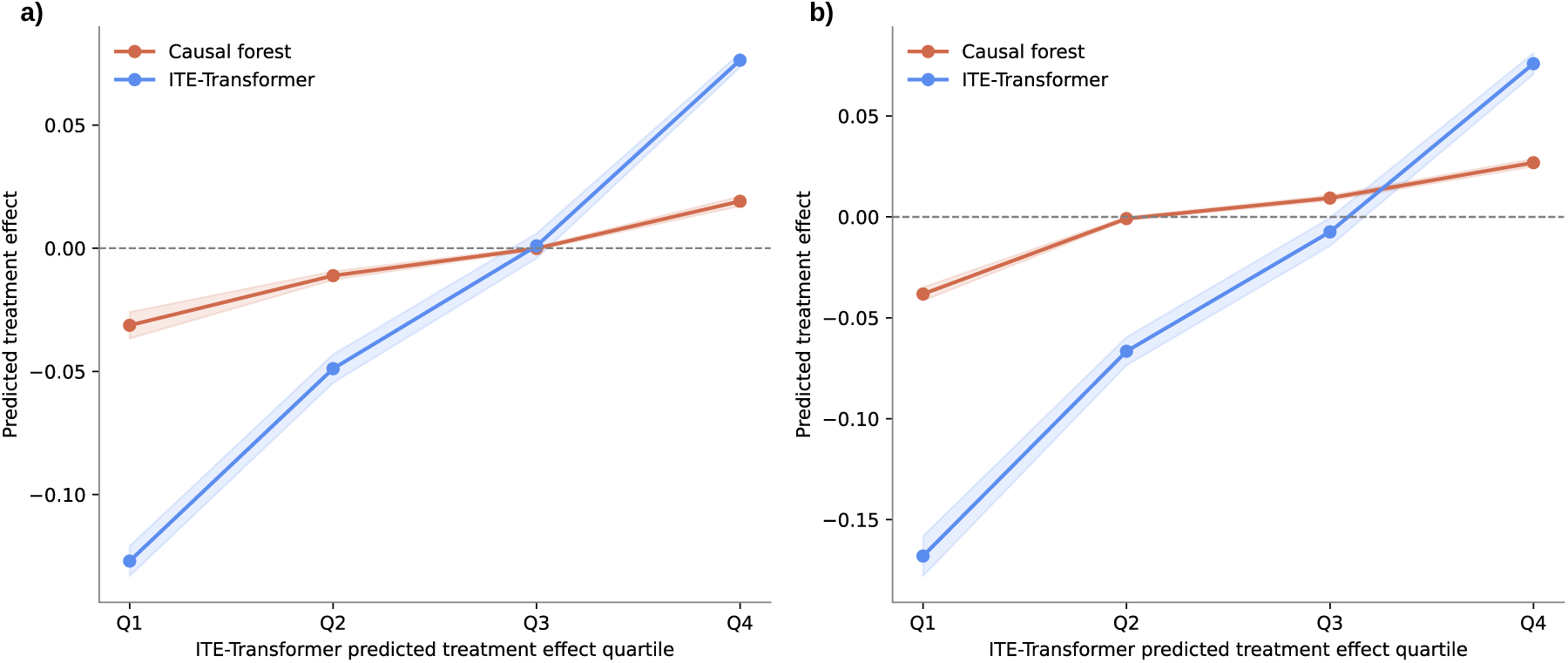
Comparison of individualized treatment effect (ITE) predictions from ITE-Transformer and causal forests. We trained causal forest on the same pre-trained patient representations as used by ITE-Transformer to provide an alternative approach to ITE prediction. Shown is the effect score calibration. Patients were grouped according to the ITE-Transformer-predicted ITEs, and the mean predictions from both methods were compared across groups. This analysis thus assesses the agreement between two estimation approaches. **a**, GLP-1 receptor agonists vs DPP-4 inhibitors task. **b**, Immunotherapy vs chemotherapy task. The broad agreement in the group-level predictions and their ranking supports the robustness of the treatment effect patterns identified by ITE-Transformer.

### Hyperparameters for the ITE-Transformer

**Table S9:** Hyperparameters used for pretraining the ITE-Transformer.

| **Hyperparameter** | **Value** |
|---|---|
| Layers | 6 |
| Heads | 4 |
| Attention heads | 4 |
| Feed-forward expansion multiplier | 4 |
| Embedding dimension | 128 |
| Minimum sequence length | 20 |
| Maximum sequence length | 128 |
| Total number of parameters | 2,570,693 |
| Batch size | 128 |
| Learning rate scheduling | Cosine annealing with linear warmup |
| Training steps | ∼110k |
| Warmup steps | ∼8k |
| Maximum learning rate | 5e-4 |
| Dropout | 0.1 |
| Weight decay | 0.01 |
| Gradient clipping | maximum norm = 1.0 |
| Training fraction | 0.85 |
| Validation fraction | 0.15 |
| Loss function | Cross entropy |
| Selection metric | Validation loss |
| Random seeds | [17, 24, 51, 103, 204] |

**Table S10:** Hyperparameters used for fine-tuning the ITE-Transformer. Stage 1: nuisance models. The same hyperparameters are used in both downstream tasks.

| **Hyperparameter** | **Value** |
|---|---|
| Number of cross-fitting folds | 5 |
| Validation fraction per-fold | 0.15 |
| Freeze transformer weights | false |
| Epochs | 50 |
| Batch size | 64 |
| Head dropout | 0.2 |
| Transformer dropout | 0.3 |
| Learning rate scheduling | linear decay |
| Maximum learning rate for head weights | 1e-4 |
| Maximum learning rate for transformer weights | 5e-6 |
| Minimum learning rate for head weights | 0 |
| Minimum learning rate for transformer weights | 0 |
| Weight decay | 0.1 |
| Selection metric | Validation binary log loss |
| Loss | see Equation 6 |
| Outcome loss weight *λ*<sub>*μ*</sub> | 1 |
| Random seeds | [17, 24, 51, 103, 204] |

**Table S11:** Hyperparameters used in fine-tuning ITE-Transformer. Stage 2: CATE learner.

| **Hyperparameter** | **Value** |
|---|---|
| Freeze transformer weights | true |
| Epochs | 30 |
| Batch size | 64 |
| Dropout | 0.3 |
| Learning rate scheduling | linear decay |
| Maximum learning rate for head weights | 5e-5 |
| Minimum learning rate for head weights | 0 |
| Weight decay | 0.1 |
| Loss | huber loss |
| Random seeds | [17, 24, 51, 103, 204] |

### Hyperparameters of the baselines

**Table S12:** Hyperparameters for the NP-Transformer.

| **Hyperparameter** | **Value** |
|---|---|
| Layers | 6 |
| Heads | 4 |
| Attention heads | 4 |
| Feed-forward expansion multiplier | 4 |
| Embedding dimension | 128 |
| Minimum context length | 20 |
| Maximum context length | 128 |
| Epochs | 60 |
| Batch size | 64 |
| Head dropout | 0.2 |
| Transformer dropout | 0.3 |
| Learning rate scheduling | linear decay |
| Maximum learning rate head weights | 1e-4 |
| Maximum learning rate transformer weights | 5e-6 |
| Minimum learning rate head weights | 0 |
| Minimum learning rate transformer weights | 0 |
| Weight decay | 0.1 |
| Selection metric | Validation binary log loss |
| Loss | see Equation 6 |
| Outcome loss weight *λ*<sub>*μ*</sub> | 1 |
| Random seeds | [17, 24, 51, 103, 204] |

**Table S13:** Hyperparameters for the random forest baseline. Separate models are trained for propensity score and outcome prediction. Propensity and outcome models share the same hyper-parameters.

| **Hyperparameter** | **Value** |
|---|---|
| Top history tokens | 512 |
| Number of trees | 500 |
| Maximum-depth options | 8, 16, 24 |
| Minimum-samples-per-leaf options | 1, 10, 25 |
| Maximum features | Square root of the total number of input features (scikit-learn option sqrt) |
| Selection metric | Validation binary log loss |
| Random seeds | [17, 24, 51, 103, 204] |

**Table S14:** Hyperparameters for the XGBoost baseline. Separate models are trained for propensity score estimation and outcome prediction. Propensity and outcome models share the same hyperparameters.

| **Hyperparameter** | **Value** |
|---|---|
| Top history tokens | 512 |
| Number of estimators | 500 |
| Learning rate | 0.05 |
| Maximum-depth options | 3, 6, 9 |
| Minimum-child-weight options | 5.0, 8.0 |
| Subsample fraction | 0.80 |
| Column-sampling fraction | 0.80 |
| *L*<sub>2</sub> regularization strength | 1.0 |
| Tree method | hist |
| Selection metric | Validation binary log loss |
| Random seeds | [17, 24, 51, 103, 204] |

## Notes

### Competing Interest Statement

The authors have declared no competing interest.

### Author Declarations

No ethical approval was necessary at the host institution (LMU Munich; LMU SOM) due to the de-identified, non-interventional design of the study. The original data collection for the NIH All of Us Research Program received ethical approval from its Institutional Review Board (IRB); see https://www.nih.gov/allofus/about/who-we-are/all-of-us-institutional-review-board

